# Efficient and Secure *μ*-Training and *μ*-Fine-Tuning for Edge-Based TinyML with Future-Guided Self-Distillation

**DOI:** 10.1101/2025.01.30.25321374

**Authors:** Zhaojing Huang, Leping Yu, Luis Fernando Herbozo Contreras, Omid Kavehei

## Abstract

This study presents a novel, computationally efficient training framework demonstrated through bio-signal processing on edge medical devices. The approach integrates conventional full training with an innovative *µ*-Training technique, wherein the encoder and decoder of a compact model remain frozen while only the middle layer is updated. This design is further enhanced by a novel *Future-Guided Self-Distillation* mechanism that leverages the model’s anticipated future state in training to boost performance and improve generalization on unseen data, using electrocardiogram (ECG) signals as the primary case study. Additionally, *µ*-Fine-Tuning facilitates on-device adaptation under resource-constrained conditions. We validate our framework using in-sample data from the Telehealth Network of Minas Gerais (TNMG) and out-of-sample testing on the China Physiological Signal Challenge 2018 (CPSC) datasets. Experimental results demonstrate that our integrated strategy (combining full training, self-distilled *µ*-Training, and *µ*-Fine-Tuning) consistently matches or surpasses conventional methods while significantly improving computational efficiency and mitigating catastrophic forgetting. Deployment on Radxa Zero hardware underscores the approach’s practical applicability and scalability. Moreover, a demonstration incorporating the proposed self-distilled *µ*-Training into standard training procedures reveals performance improvements. This highlights the technique’s potential for broader applications beyond medical diagnostics and TinyML systems, paving the way for its integration into existing training mechanisms to elevate overall model performance.

## 1 Introduction

One of the key challenges in the medical domain is developing models that can be personalized for individual patients. To achieve this, models need to adapt to new personal data. However, when it comes to edge devices, their often severely limited computational and memory capacity makes local fine-tuning on new data impractical.

Traditionally, this challenge is addressed by transferring data to external servers for processing [1]. This raises significant data security concerns, as such transfers are vulnerable to hacking and unauthorized access [2].

An alternative approach is Federated Learning (FL), which exchanges gradients instead of raw data to mitigate privacy risks [3]. However, recent studies have shown that even gradients can be exploited to reconstruct sensitive data [4, 5]. Moreover, transferring either data or gradients can rapidly drain the device’s battery (if battery-powered), as data transmission, which often has to be wireless, consumes substantial energy [6].

Our previous work introduced a novel method for finetuning large foundational models without requiring data or gradient transfer [7]. This approach facilitates finetuning large models directly on resource-limited devices, enhancing data security and efficiency. However, recent advancements in machine learning have led to the development of smaller models that can deliver performance comparable to larger models [8]. Despite their smaller model size, fine-tuning these models on resource-constrained edge devices poses a significant challenge. Huang et al. [7]’s *µ*-Trainer, initially designed for finetuning larger models, offers valuable insights into improving the efficiency of fine-tuning smaller models. This approach allows for more efficient fine-tuning in resource-constrained environments. A key concept in *µ*-Trainer is to focus knowledge condensation in specific model layers, freeing up other layers to better adapt to new information. Building on this idea, we aim to develop a new methodology tailored for fine-tuning smaller models for personalized use, emphasizing maximizing resource efficiency. The knowledge distillation approach employed in the previous study [7] enhanced performance. Building on that insight, we developed a novel self-distillation technique—eliminating the need for an external teacher model—that, when integrated with the proposed *µ*-Training process, yields promising results and shows potential for broader applications. To demonstrate the effectiveness of this approach, we utilized one of our previously developed tiny models on electrocardiogram (ECG) data [8], highlighting its potential for medical applications.

ECG is a critical diagnostic tool for assessing cardiac activity, providing information about the electrical signals generated by the heart muscle during heart movement. It plays a pivotal role in detecting cardiac irregularities, aiding in the early diagnosis of heart conditions, and mitigating the risk of stroke [9]. The challenges of resource-constrained environments, data privacy, and security are particularly pressing in the AI-driven biomedical field. Striking a balance between leveraging data for medical advancements and safeguarding individual privacy is crucial. Addressing these challenges requires a multidisciplinary approach, combining several domains of expertise such as AI, healthcare system processes, and legal frameworks to mitigate privacy risks and serious consequences for patients, staff or entities involved [10]. Our proposed method addresses this critical issue, as envisioned in Fig. 1.

**Figure 1:**
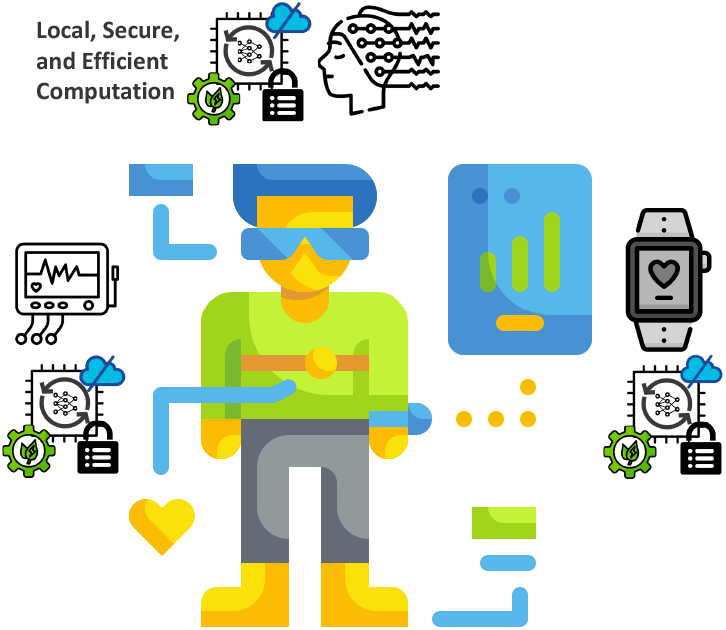
The vision of this work is to develop a method for training and fine-tuning small yet accurate machine learning models on personal data, enabling secure model personalization while enhancing efficiency for on-device fine-tuning. This approach is particularly crucial in the medical domain, where data security and resource efficiency are paramount.

With the proliferation of Internet of Things (IoT) applications and the potential integration with machine learning at the edge, the need for personalization at the edge is expected to grow. Integrating IoT and edge-based Artificial Intelligence (AI) offers compelling benefits, such as ultra-low latency, strengthened data security, enhanced privacy protections, and significant cost reductions [11]. This study focuses on a medical application case and aims to expand its application beyond the medical domain to other fields of IoT.

The key novel contributions of the proposed methods are as follows:

- **Innovative** *µ***-Training &** *µ***-Fine-tuning** — This approach introduces a unique training method that freezes the encoder and decoder while making only the middle layer trainable, allowing layer-focused training to enhance model performance.
- **Novel Future-Guided Self-Distillation** — A novel self-distillation approach leverages the model’s anticipated future state to guide training, enhancing performance and generalization. This technique is integrated into *µ*-Training to boost model efficacy further.
- **Computational Efficiency** — The proposed technique achieves results on par with or superior to traditional training while significantly reducing computational overhead.
- **Edge Deployment** — The technique is validated on ECG data and deployed on devices like Radxa Zero, demonstrating strong potential for real-life applications in resource-constrained environments.
- **Integrated Self-Distilled** *µ***-Training** — Self-distilled *µ*-Training is integrated into the standard training procedure, demonstrating its superiority over conventional training methods and its potential for future adoption in regular model training.

### 1.1 Background

Building on the *µ*-Trainer framework [7], we introduce tailored *µ*-Training and *µ*-Fine-Tuning strategies for tiny models, complemented by a novel self-distillation approach. This new method extends the core idea of *µ*-Trainer by enabling training and fine-tuning to focus on specific model parts, optimizing computational and memory efficiency. Additionally, this approach aims to enhance the performance of models trained and fine-tuned using this methodology.

#### 1.1.1 Tiny Machine Learning (TinyML)

Our previous works investigated the potential of tiny models that achieve strong performance while maintaining a compact size in terms of weights and parameters. We identified the Diagonal State Space Sequence (S4D) model as highly effective for processing sequential data [12, 13]. Additionally, we explored the capabilities of specialized neurons, such as Neural Circuit Policies (NCP) [14, 15, 16] and Kolmogorov-Arnold Networks (KAN) [17, 18], both of which demonstrated promising results. Building on these findings, we successfully integrated the S4D model with NCP neurons to create an extremely compact model that delivers strong performance on timeseries data [8]. For this study, we will utilize this compact, lightweight model to demonstrate the effectiveness of our proposed methodology. The model can be divided into three (3) primary layers: the encoder, the S4D layer, and the decoder. Further details about the model’s architecture will be provided in Section 2.1.

#### 1.1.2 Privacy and Data Security

Several approaches, such as FL and server-based data upload, have been developed for fine-tuning models [19]. FL has been widely adopted in fields like electronic health record management, medical image processing, and remote health monitoring [3].

However, risks of user data leakage remain. Zhu et al. [4]introduced Deep Leakage from Gradients (DLG), a method capable of reconstructing training inputs and labels directly from gradients. Similarly, Boenisch et al. [5]demonstrated vulnerabilities in FL, showing that even when protected by Distributed Differential Privacy (DDP) and Secure Aggregation (SA), FL remains susceptible to breaches. These findings highlight that FL, despite its advancements, does not fully guarantee user privacy. Therefore, innovative methods prioritizing user privacy and robust data security are essential for advancing edge AI systems.

#### 1.1.3 Personalized Model Adaptation

Personalization based on specific user data is essential to enhance the suitability of models for individual users. However, transmitting data outside the device is energy-inefficient, and combined with privacy and data security concerns, on-device fine-tuning emerges as a feasible solution. These challenges highlight the need for local, on-device training solutions [20].

### 1.2 Overview of the Proposed Approach

The proposed method, outlined in Fig. 2, is structured into three steps to fine-tune a small model for more personalized usage while maintaining user privacy. Personal data is retained on the device throughout this process, mitigating potential privacy concerns. At no stage is information such as input data batches *x*, true labels *y*, or gradients ∇*W* transferred beyond the device. Finetuning is performed locally using privately collected personal data, and the iterative processes for each step are described as follows.

**Figure 2:**
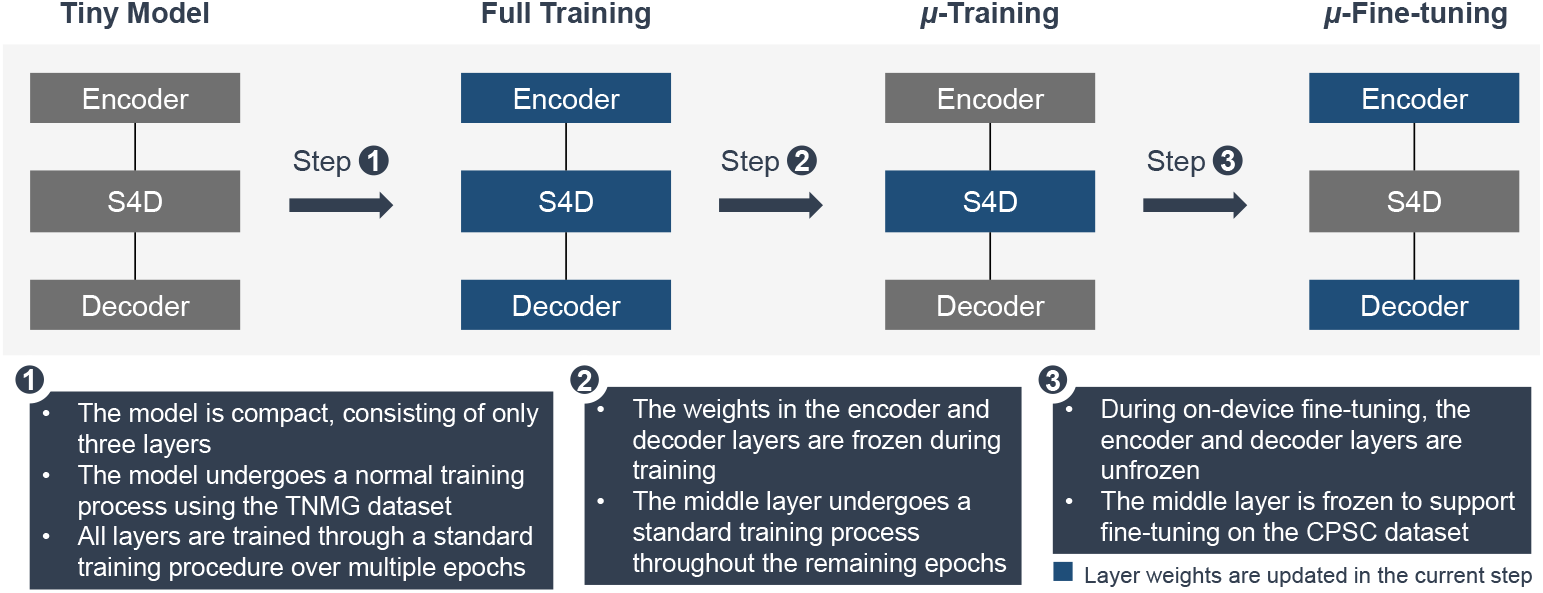
The chart provides a detailed overview of the methodology, divided into three distinct steps. Step 1 - Full Training: In this step, the model operates in full training mode, where all weights are updated through the standard training process. Step 2 - *µ*-Training: During *µ*-Training, the weights in the encoder and decoder are frozen, while the middle S4D layer remains trainable. Step 3 - *µ*-Fine-Tuning: In the final step, the trained model undergoes *µ*-Fine-Tuning on the device. Here, the middle S4D layer is frozen while the encoder and decoder are set to be trainable, enabling the model to adapt its weights to new, unseen data. Green highlights in the chart indicate the layers where weights are updated during each respective step.

The methodology is further detailed in Section 3. Two datasets were employed to simulate the experimental process while ensuring minimal use of personal data. The first dataset is the original training data for pre-training the model. In contrast, the second dataset is specifically designed to evaluate the effectiveness of the proposed fine-tuning approach. Comprehensive descriptions of these datasets, including their characteristics and relevance to the experiments, are provided in Section 4.

To further enhance the performance of the proposed technique and build upon our previous work on knowledge distillation [7], we have developed a novel future-guided self-distillation approach that integrates into the *µ*-Training process displayed in Fig. 3. This integration improves model performance and extends the applicability of self-distilled *µ*-Training beyond the medical and TinyML domains, demonstrating promising potential for advancing standard model training methodologies. A thorough and detailed description of the proposed self-distillation method is presented in Section 3.7.

**Figure 3:**
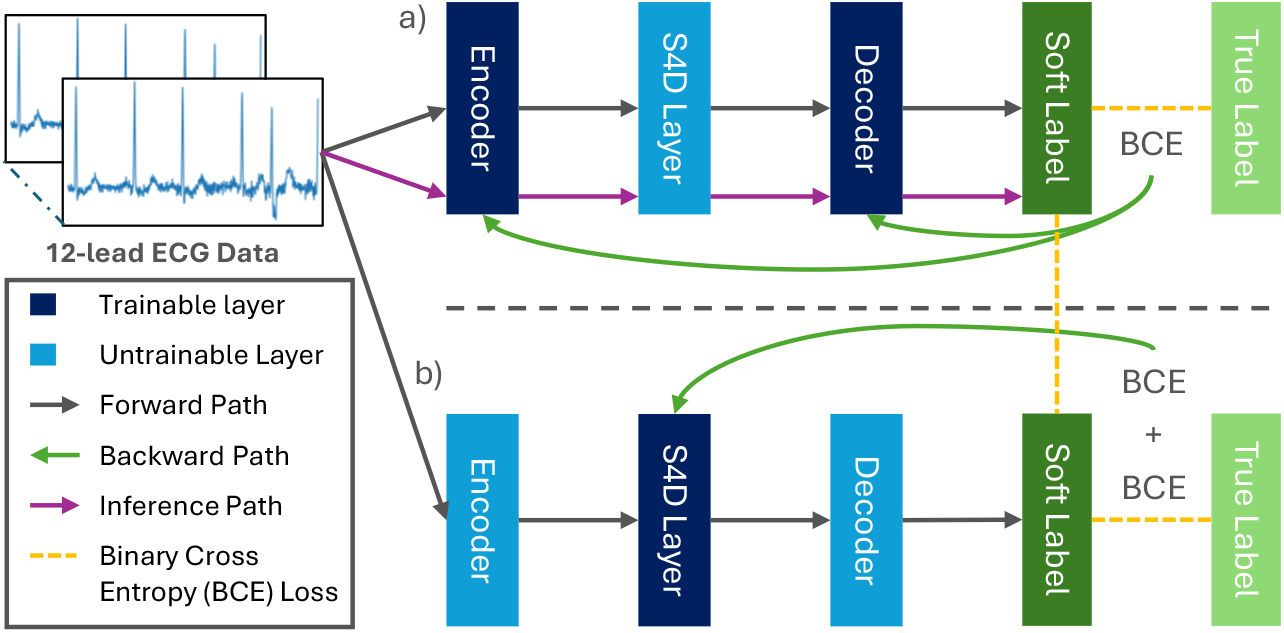
This figure illustrates one iteration of the Integrated Future-Guided Self-Distilled *µ*-Training process. a) Demonstrates the first training phase: i) The model is a cloned version with a frozen middle layer, while the encoder and decoder remain trainable. ii) The model performs a forward pass, computing the Binary Cross-Entropy (BCE) loss against the true labels. iii) The unfrozen layers (encoder and decoder) are updated based on the computed loss on the backward path. iv) The model generates soft labels through inference. b) Depicts the second training phase under the *µ*-Training configuration: v) The real model is activated, with the middle layer unfrozen while the encoder and decoder layers remain frozen. vi) The model performs a forward pass to generate soft labels. vii) An integrated BCE loss is computed with both the true labels and the soft labels generated in the previous phase. viii) A backward pass updates the model’s S4D layer, refining its feature representations.

Compared to conventional methods, where data is sent outside the device for external fine-tuning, and Federated Learning (FL), where gradients are shared outside the device for global aggregation, our proposed method significantly advances privacy protection, personalization, and on-device training. A qualitative comparison is provided in Table 1, which underscores the unique contributions of this work in comparison with other approaches that share the same aim.

**Table 1:** Comparison of various methodologies (✓: Yes; *×*: No; –: Challenging)

|  | Privacy | Personalize | On-device Learning | Compute Efficiency |
| --- | --- | --- | --- | --- |
| Conventional | × <sup>†</sup> | ✓ | $-$ <sup>‡</sup> | × |
| FL | × [4] | $-$ [5] | $-$ | × |
| Proposed | ✓ | ✓ | ✓ | ✓ |

<sup>†</sup> Transmitting user data externally presents substantial privacy risks, as it exposes sensitive information to potential unauthorized access by malicious actors, such as hackers, who could exploit this direct access.
<sup>‡</sup> Edge devices typically lack the necessary memory and computational power for full model training or fine-tuning, a standard practice in conventional methods.

## 2 Related Works

### 2.1 Tiny Diagonal State Space Model

The Tiny Diagonal State Space Model (TDSSM) is a simplified and computationally efficient approach to modeling dynamical systems [8]. The model builds on the standard state-space framework, which describes how the system evolves and how the internal state relates to observed measurements [8]. In the TDSSM, these equations are streamlined to minimize computational and memory demands, making the model suitable for edge devices and real-time applications [8].

One defining characteristic of S4D is using diagonal matrices for key components such as the state transition or observation matrices [12]. This diagonal assumption greatly reduces computational complexity as operations like matrix multiplications and inversions become simpler and more efficient. While diagonalization limits the model’s expressiveness compared to general state-space models, it is sufficient for capturing essential patterns in many practical applications [8]. The TDSSM model has excellent performance on large amounts of data, which is suitable for complex datasets [12, 13]. The Neural Circuit Policy (NCP) decoder is utilized in this model. It draws inspiration from the nervous system of the nematode Caenorhabditis elegans, which features a highly efficient network of neurons that facilitate enhanced communication and coordination. This design enables the NCP decoder to achieve efficient and robust processing capabilities [14, 15, 16]. The complete model structure is shown in Fig. 4.

**Figure 4:**
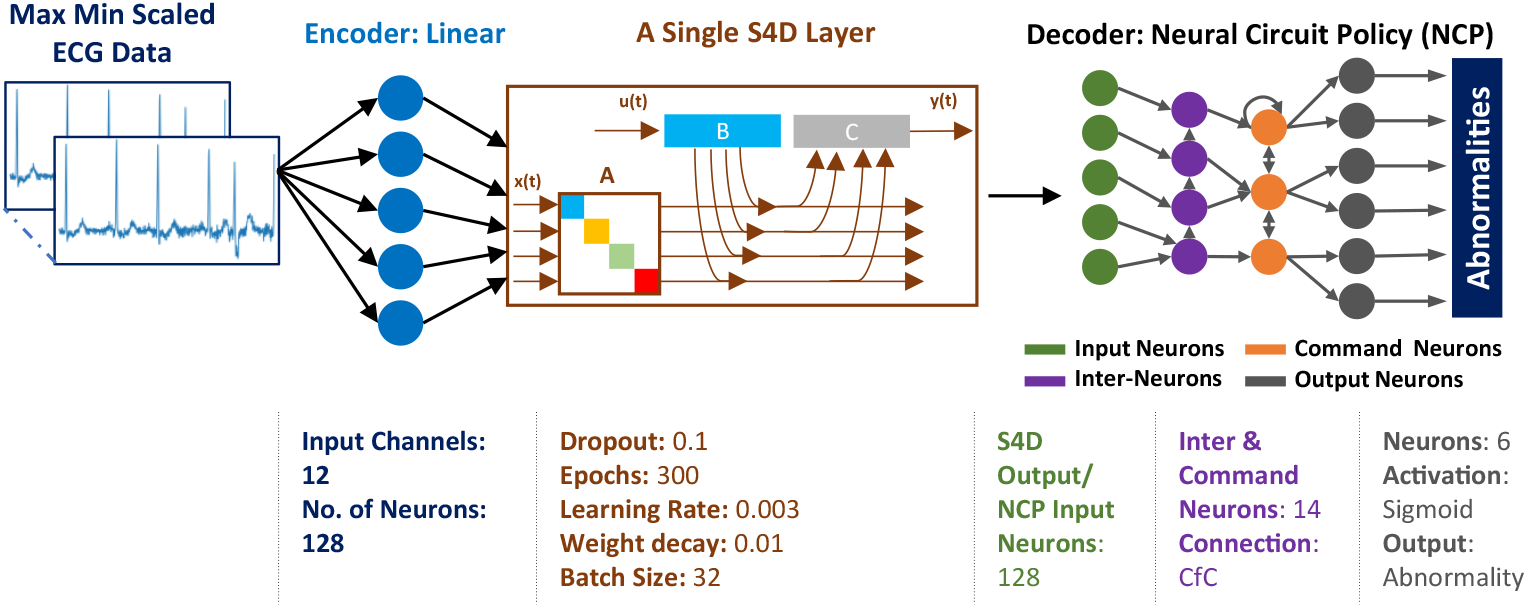
Huang et al. [8]’s Tiny Diagonal State Space Model features a single linear encoder, single S4D layer, and simple NCP neurons serving as the decoder. Adapted from Huang, Z., et al. APL Mach. Learn. 2, 026112 (2024) [8]. https://doi.org/10.1063/5.0191574. Licensed under CC BY 4.0.

For cardiac abnormality detection, as explored in the referenced paper, the TDSSM provides an efficient way to model the dynamics of cardiac signals, such as ECGs [8]. The approach can detect abnormalities potentially on low-power wearable devices by leveraging the model’s simplicity and low computational footprint. This makes it a promising tool for portable health monitoring systems [8].

### 2.2 µ-Trainer

*µ*-Trainer is a novel method that was developed to enable the training of large-scale machine learning models on resource-constrained devices. This method utilizes knowledge distillation, which is performed using a “student” model trained to replicate the behavior and knowledge of a larger, more complex “teacher” model. By focusing on efficient knowledge transfer, *µ*-Trainer ensures that even resource-limited devices can benefit from the capabilities of large machine learning models [7].

One of the key features of *µ*-Trainer is the ability to compress knowledge. The method selectively transfers the most relevant information from the teacher model to the student model while eliminating redundant or unnecessary knowledge. This approach allows the student model to achieve performance levels close to the teacher model but with a significantly smaller computational and memory footprint [7]. Utilizing the *µ*-Trainer for finetuning ensures that the data and gradients are not communicated outside the device, thereby preserving privacy [4, 5].

To optimize training for edge devices, *µ*-Trainer utilized dynamic layer freezing, a strategy where less critical layers of the model are frozen during training. This reduces computational demands without significantly affecting the model’s accuracy, making it particularly suited for edge applications [7].

Another distinctive feature of *µ*-Trainer is its use of progressive knowledge transfer. This training strategy involves transferring knowledge from the student model to the larger one and creating a new student model to adapt to newer data. This process is repeated in a loop, enabling continuous improvement of the larger model over time [7].

### 2.3 µ-Training and µ-Fine-tuning Tiny Models

The *µ*-Trainer was initially designed to enhance the security of fine-tuning large foundational models locally within resource-constrained environments. However, its underlying concept could also benefit the personalization of smaller models in the medical domain, an area of critical importance. Beyond ensuring the security of personal data, the *µ*-Trainer can potentially improve the efficiency of model personalization and fine-tuning processes. The core idea of the *µ*-Trainer is to enable one part of the model to focus on learning specific data features while allowing another part of the model greater flexibility to adapt to new data. This study explores how these benefits can be applied to fine-tune a smaller model (TDSSM) we previously developed and tested. We have designed the technique to enable the small model to undergo a standard training process, followed by *µ*-Training and *µ*-Fine-Tuning on new data, to achieve effective personalization.

### 2.4 Self-distillation

Furlanello et al. [21] explored the performance of Knowledge Distillation by using the same model for both student and teacher model, providing the technology background for the self-distillation.

Self-distillation is a method in which a model is trained using its predictions as the target, bypassing the need for external teacher models [22]. This process allows the model to refine its knowledge and improve performance, effectively “learning from itself.” While it shares similarities with knowledge distillation, self-distillation can help enhance generalization, reduce overfitting, and improve robustness in tasks like classification and regression, as the model continually learns to predict better outputs from its previous predictions [22].

However, current self-distillation uses a model’s soft predictions as teacher signals to refine training and improve generalization. In contrast, our method integrates with *µ*-Training, guiding the model with predictions based on its projected future weights, enhancing performance without adopting them.

## 3 Methods

To evaluate the feasibility of our proposed technique, we designed a comprehensive experimental methodology as outlined in Fig. 2. This process is divided into three key steps.

In Step 1, the model undergoes full training on an external server, where all weights are updated through the standard backpropagation process. The weights are adjusted using the Eq. 1.

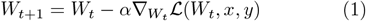

where *W*_*t*_ represents the model weights at iteration *t, α* is the learning rate, 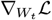 denotes the gradient of the loss function ℒ, and *x, y* are the input data and labels, respectively.

In Step 2, during *µ*-Training, the encoder and decoder layers are frozen, and only the middle S4D layer remains trainable. The updated weights of the S4D layer are computed as Eq. 2.

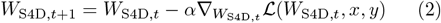

Here, *W*_S4D,*t*_ denotes the weights of the trainable S4D layer, while the encoder and decoder weights remain fixed. We propose a novel self-distillation method that enhances our overall approach and seamlessly integrates with the *µ*-Training component. A dedicated section is provided to explain and empirically evaluate the proposed distillation technique.

In Step 3, during *µ*-Fine-Tuning, the middle S4D layer is frozen, and the encoder and decoder layers become trainable to allow the model to adapt to new, unseen data. The weights are updated using Eq. 3.

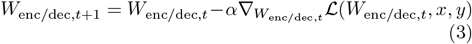

where *W*_enc/dec,*t*_ represents the trainable weights of the encoder and decoder layers during fine-tuning. Detailed execution of the proposed *µ*-Training method is summarized in Algorithm 1.

The following subsections will describe the detailed implementation of each step in the proposed methodology. In addition, we conducted supplementary experiments to broaden our insights and further refine our approach. Specifically, we began by investigating the impact of the positioning of *µ*-Training. We will also explore the interaction between the proposed method and model size and examine the integration of our novel self-distillation technique with *µ*-Training after demonstrating applications of *µ*-Training and *µ*-Fine-tuning strategy.

### 3.1 µ-Training Positioning

To examine the effect of sequencing full training and *µ*-Training within the overall training process, we conducted preliminary experiments. We compared the performance of models trained with *µ*-Training applied at various stages. We applied *µ*-fine-tuning to these models using a separate generalization dataset from another source, simulating a personal data scenario, and evaluated their performance on both the training and generalization datasets. The resulting performance differences were then analyzed. Regardless of the order, the model undergoes full training, *µ*-Training, and *µ*-Fine-Tuning. We utilized the TDSSM model, adapted from the work of Huang et al. [8], following the same training process outlined in Sections 3.2, 3.3, and 3.4, which includes full training, *µ*-Training, and *µ*-Fine-Tuning. The only difference lies in the positioning of *µ*-Training within the overall training process.

#### Algorithm 1

*µ*-Training and *µ*-Fine-Tuning Process

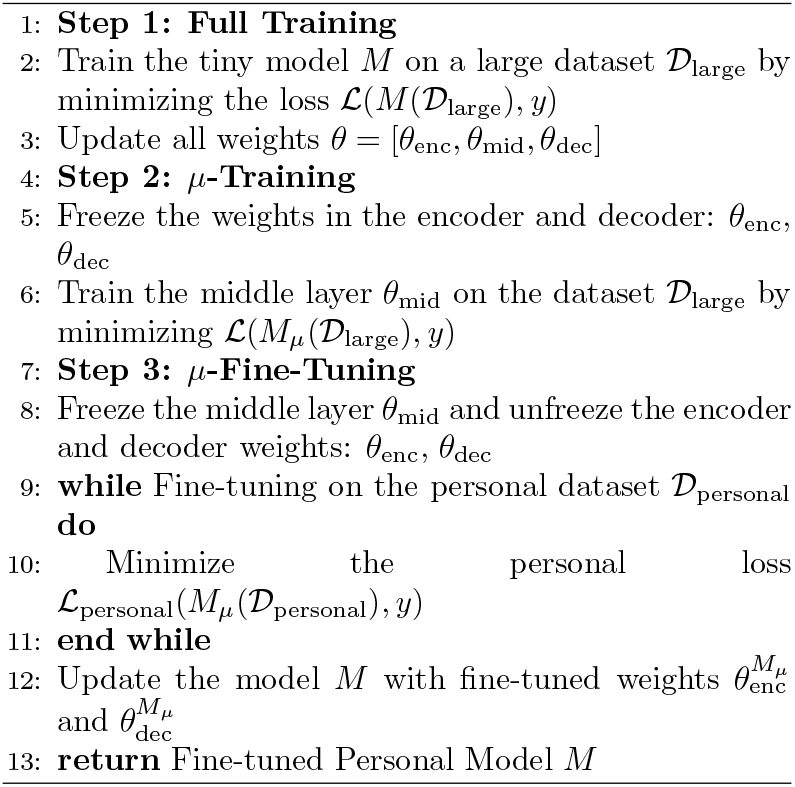

### 3.2 Step 1 - Full Training

The first step involves training the TDSSM, adapted from the work of Huang et al. [8].

This highly efficient model features just 58,509 parameters and a compact size of only 242 KB. Despite its minimal footprint, it delivers performance on par with significantly larger foundational models [8]. The training dataset is utilized during the training phase, with the ECG data subjected to min-max scaling to normalize its range, thereby improving the model’s generalizability and performance [8].

The model was trained on a Tesla V100S-PCIE-32GB GPU, using under 3 GB of memory and completing within a day. Key hyperparameters, such as a batch size of 32, 300 epochs, and a learning rate of 0.003, were determined based on previous experiments in [8] to ensure stability and strong performance. The Adam optimizer and binary cross-entropy loss function were selected for efficient multi-label classification. A Cosine Annealing Learning Rate Scheduler was implemented to adjust the learning rate smoothly, preventing instability and ensuring steady convergence. These configurations proved effective in earlier work in [8]. The model design was selected based on our previous work [12, 8], where we conducted ablation studies on stacked S4D layers [12] to assess the model’s performance concerning the number of layers. Our findings indicate that many layers are not necessary for optimal performance.

The training process was divided into two phases: *Full training* and *µ-Training*. All model layers and weights were trained in a standard manner during full training. In contrast, *µ*-Training involved a more targeted training strategy. Various combinations of full training and *µ*-Training were explored to evaluate the performance of the proposed technique. Notably, the total number of epochs for both phases combined was maintained at 300. By keeping the total number of epochs constant, we can isolate and analyze the specific impact of *µ*-Training.

### 3.3 Step 2 - *µ*-Training

The second step focuses on *µ*-Training, a specialized phase designed to refine the model while preserving the integrity of specific components. During *µ*-Training, the weights in the encoder and decoder layers are frozen, meaning these layers retain their pre-trained parameters from the previous step without further updates. Instead, all training updates are concentrated solely on the middle layer, which, in this case, is the S4D layer, comprising 49,920 weights out of the total 58,509. The *µ*-Training process is conducted on the training dataset, also utilized during the full training phase.

This targeted approach allows the S4D layer to adapt more effectively to the training data while maintaining the structural stability provided by the frozen layers. By limiting updates to a single layer, *µ*-Training reduces computational overhead and enhances the model’s ability to generalize without overfitting.

The entire training process, including full training and *µ*-Training, is designed to conclude at 300 epochs. *µ*-Training occupies the remaining epochs after the initial full training phase, ensuring a seamless transition from broad training across all layers to a more focused refinement of the S4D layer. This methodical division aims to optimize the model’s performance, balancing efficiency and accuracy. To determine the most efficient balance between full training and *µ*-Training, we varied the number of *µ*-Training epochs from 50 to 300 within a total of 300 training epochs, using intervals of 50 to observe the changes.

### 3.4 Step 3 - *µ*-Fine-tuning

After completing the full training and *µ*-Training phases, the model undergoes the *µ*-Fine-Tuning process. Unlike the *µ*-Training phase, where only the middle S4D layer was updated, *µ*-Fine-Tuning freezes the S4D layer and allows only the encoder and decoder layers to adjust their weights. This strategic adjustment enables the model to adapt to new data while preserving the specialized functionality of the S4D layer. The encoder and decoder layers contain 8,589 weights, which account for approximately 14.68% of the total weights.

The fine-tuning process is limited to 50 epochs to achieve rapid adaptation and computational efficiency. This phase utilizes a small, balanced subset of the generalization dataset, a distinct dataset from the original training data, to simulate real-world scenarios where the model must adapt to new, personalized data. Such an approach reflects practical applications where models are fine-tuned on limited data to personalize their performance effectively. Using a balanced dataset has been demonstrated to enhance personalization performance [7], making it an ideal approach for optimizing the model’s effectiveness in individual applications. As the generalization and training datasets may contain different classes of abnormalities, the loss function must be redesigned to account for these differences. This ensures that irrelevant classes do not interfere with the finetuning results.

### 3.5 Edge Deployment of *µ*-Fine-tuning

To verify the effectiveness of the proposed technique, we deployed the model on the Radxa Zero, a compact single-board computer powered by the Amlogic S905Y2 64-bit quad-core ARM Cortex-A53 processor (1.8 GHz) with 4 GB of LPDDR4 RAM. This resource-constrained environment is ideal for testing lightweight machine-learning models designed for edge deployment. Its compatibility with deep learning libraries like PyTorch enables efficient model development and testing, and its setup can be replicated on other edge devices, such as the Raspberry Pi, for broader applicability.

We used a model that underwent the proposed full training and *µ*-Training process, deploying it on the edge platform for *µ*-fine-tuning. This model was compared to one trained normally for 300 epochs and fine-tuned for 50 epochs on the same dataset using a GPU.

### 3.6 Impact of Model Size

To evaluate the impact of the proposed method on models with varying sizes and weight distributions across layers, we constructed several variants of the TDSSM. This was done by adjusting the model dimension parameter (*d*_model_), which controls the model’s width. In our previous experiments, we fixed *d*_model_ at 128. In this section, we vary this parameter to create model variants of different sizes.

This approach extends the insights provided by *µ*-Trainer [7], which primarily investigates the effect of model depth (i.e., the number of layers). By leveraging the flexibility of TDSSM with varying *d*_model_ values, our study shifts focus to the impact of model width. This enables a more comprehensive evaluation of the proposed methodology. Our findings provide deeper insights into the design of efficient models. They also offer valuable guidance for optimizing both the *µ*-Trainer framework and the overall model architecture discussed in [7].

### 3.7 Self-Distilled *µ*-Training

Building upon the insights from *µ*-Trainer [7], the knowledge distillation used in constructing the *µ*-Trainer has significantly enhanced both the trainer’s performance and the overall effectiveness of the method. Inspired by this, we sought to improve the proposed *µ*-Training method by exploring the possibility of incorporating knowledge distillation in this context.

#### 3.7.1 A Novel Self-Distillation Framework

Typically, knowledge distillation requires a teacher model to guide the training of a student model. However, in the case of *µ*-Training, which involves training a single tiny model, there is no separate teacher model. To address this, we propose a novel approach that allows the model to self-distill during the *µ*-Training process. Given that the top and bottom layers of the model are frozen during *µ*-Training, we construct a hypothetical configuration in which these layers are unfrozen while freezing the originally trainable layers. This is a guiding mechanism to inform the optimization of the unfrozen weights at each *µ*-Training iteration. So essentially, the model is trained based on its anticipated future of the frozen layers. Details regarding self-distillation integration are provided in Fig. 3 and its implementation in Algorithm 2.

##### Algorithm 2

One Iteration of *µ*-Training with Future-Guided Self-Distillation

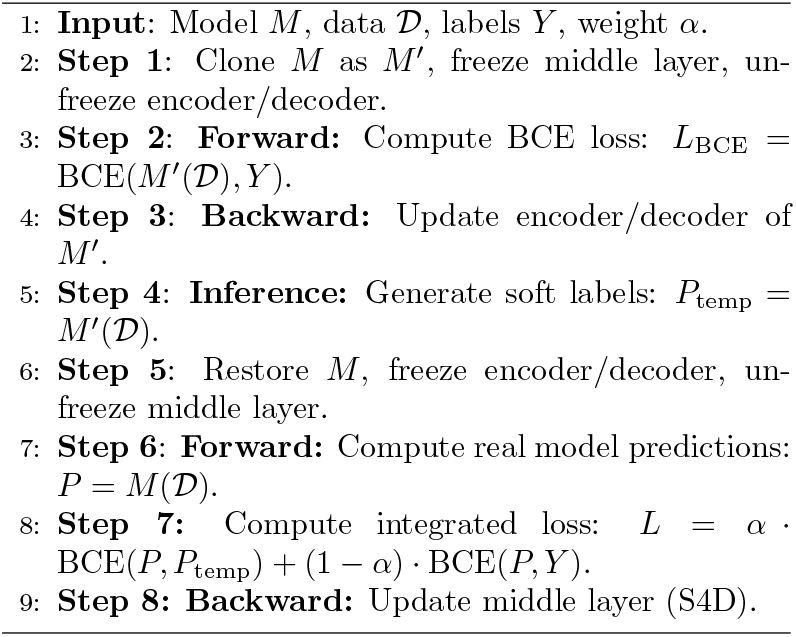

#### 3.7.2 Comparison with Existing Self-Distillation Methods

Previous approaches to Knowledge Distillation have largely relied on a two-model paradigm. Hinton et al. [23] introduced the concept where a smaller student model learns from a larger teacher model by mimicking its softened output probabilities. This technique enables model compression while maintaining performance demonstrated on MNIST and speech recognition tasks.

Romero et al. [24] extended this by introducing “hints” that are intermediate feature representations to guide the student network, improving efficiency. Traditional self-distillation techniques involve a model learning from its predictions to enhance performance. This process typically includes generating soft labels through a high-temperature softmax applied to the model’s logits, which are then used as targets for training. The model refines its decision boundaries by aligning its predictions with these soft targets, leading to improved generalization [22]. An example of self-distillation is the framework proposed by Zhang et al. [25], which divides the network into multiple sections and facilitates knowledge transfer from deeper to shallower parts within the same network. This internal knowledge transfer refines the model’s learning process, improving accuracy and generalization. In contrast, our method integrates with *µ*-Training, which allows the model to generate predictions based on its anticipated future weights, not its current state. Unlike self-distillation, where the model is guided by its past behavior, our approach leverages its projected future state as a dynamic guiding signal. Although the model never adopts these future weights due to the *µ*-Training configuration, this foresight directs the learning process during training, enabling more proactive refinement and steering the model toward better long-term performance.

## 4 Datasets

The evaluation of the proposed approach in this study was carried out using two separate datasets. The first dataset, the Telehealth Network of Minas Gerais (TNMG) dataset [26], is tailored for detecting abnormalities in both rhythm and morphology within 12-lead ECG recordings. This dataset served as the training ground for the model presented in this work. This dataset is used as the training dataset.

The second CPSC dataset was created for the China Physiological Signal Challenge 2018 [27]. It consists of labeled abnormality data for 12-lead ECG signals and was utilized during the fine-tuning stage to mimic scenarios where new and unfamiliar data are encountered. This dataset serves as a generalization dataset, simulating a personal dataset to evaluate the proposed technique’s effectiveness in personalization scenarios.

### 4.1 TNMG

The TNMG dataset consists of 2,322,513 distinct labeled samples, each representing 10 seconds of 12-lead ECG data. These samples cover six different types of abnormalities: Atrial Fibrillation (AF), First Degree Atrioventricular Block (1dAVb), Left Bundle Branch Block (LBBB), Right Bundle Branch Block (RBBB), Sinus Bradycardia (SB), and Sinus Tachycardia (ST) [26]. The sampling rate for this dataset is 40 Hz.

A balanced subset was created by randomly selecting 3,000 samples for each of the six abnormality types and an additional 3,000 samples representing healthy ECG readings, resulting in 21,000 samples for model training. In cases where patients exhibited multiple abnormalities, any additional samples required to reach the subset size of 21,000 were randomly chosen from the full TNMG dataset. Further details on these abnormalities can be found in Table 2.

**Table 2:** Classification of Abnormalities in the TNMG Dataset with Descriptions.

| Abnormality | Description |
| --- | --- |
| Atrial Fibrillation (AF) | A common arrhythmia characterized by rapid and irregular heartbeats, leading to an irregular ventricular response [28]. |
| Sinus Tachycardia (ST) | A condition where the heart rate exceeds 100 beats per minute, originating from the sinoatrial node. It can result from various physiological or pathological conditions [29]. |
| Sinus Bradycardia (SB) | Defined as a heart rate of fewer than 60 beats per minute, with impulses originating from the sinoatrial node. It can be normal in athletes or indicate underlying pathology [30]. |
| Left Bundle Branch Block (LBBB) | A delay or blockage in the left bundle branch of the heart’s conduction system, causing the left ventricle to contract later than the right [31]. |
| Right Bundle Branch Block (RBBB) | A condition where there’s a delay or blockage in the right bundle branch, leading to delayed right ventricular contraction [32]. |
| First Degree Atrioventricular Block (1dAVb) | Characterized by a prolonged PR interval exceeding 200 ms on an ECG, indicating delayed conduction through the atrioventricular node [33]. |

The dataset was standardized to a fixed length of 4,096 readings to ensure uniformity in analysis and modeling. Readings exceeding this length were truncated to streamline data processing and facilitate comparison. Fig. 5 illustrates the balanced gender distribution in the resampled dataset, enhancing generalizability and supporting robust analysis. Furthermore, the dataset reflects the age distribution of the general population, highlighting its relevance for age-related studies. Additionally, the balanced representation of different abnormalities in the dataset enhances the model’s learning efficiency and overall performance [34].

**Figure 5:**
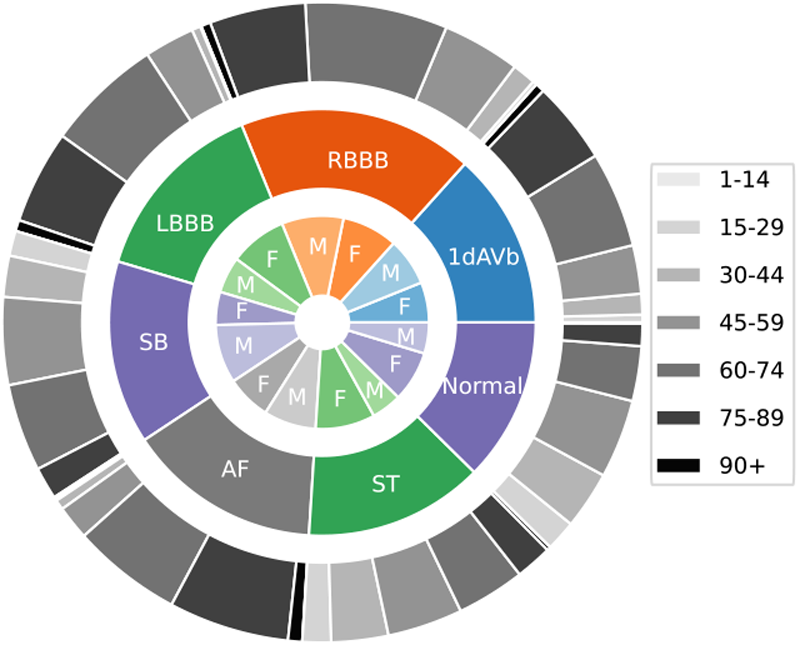
The TNMG subset is a well-balanced dataset comprising six types of abnormalities and features a higher proportion of older patients, aligning with the demographics of the general population. Adopted from Huang, Z., et al. J. of Cardiovasc. Trans. Res. (2024) [14]. https://doi.org/10.1007/s12265-024-10504-y. Licensed under CC BY 4.0.

### 4.2 CPSC

The CPSC dataset comprises 12-lead ECG recordings ranging from 6 to 60 seconds in duration, sampled at a rate of 500 Hz. To ensure compatibility with the CPSC dataset, the TNMG data was resampled to match this rate specifically for training purposes. The CPSC dataset includes ECGs from patients diagnosed with various cardiovascular conditions, accompanied by precise annotations for abnormalities. In total, the dataset covers eight distinct types of abnormalities.

This dataset, previously unused in training the model, serves as a benchmark to evaluate the fine-tuning potential of the proposed methods. While the CPSC dataset contains eight abnormality classes, four overlap with those in the TNMG dataset, making them the focus for fine-tuning evaluation. The selected abnormalities for assessment include Atrial Fibrillation (AF), Left Bundle Branch Block (LBBB), Right Bundle Branch Block (RBBB), and First Degree Atrioventricular Block (1dAVb). The remaining abnormalities—Premature Atrial Contraction (PAC), Premature Ventricular Contraction (PVC), ST-segment Depression (STD), and ST-segment Elevation (STE)—were excluded from this study.

As part of data preparation, entries with missing values were removed, resulting in a final dataset of 6,877 unique ECG tracings. All recordings were standardized to 4,096 readings for consistency, with excess data truncated during the cleaning process. Fig. 6 illustrates the distribution of abnormalities within the CPSC dataset. Analysis of the CPSC dataset revealed a gender imbalance, with a higher proportion of male patients than females. However, the age distribution aligns closely with the general population, showing a higher representation of older individuals. While most abnormalities are evenly represented, a slight under-representation of LBBB was observed compared to other abnormalities.

**Figure 6:**
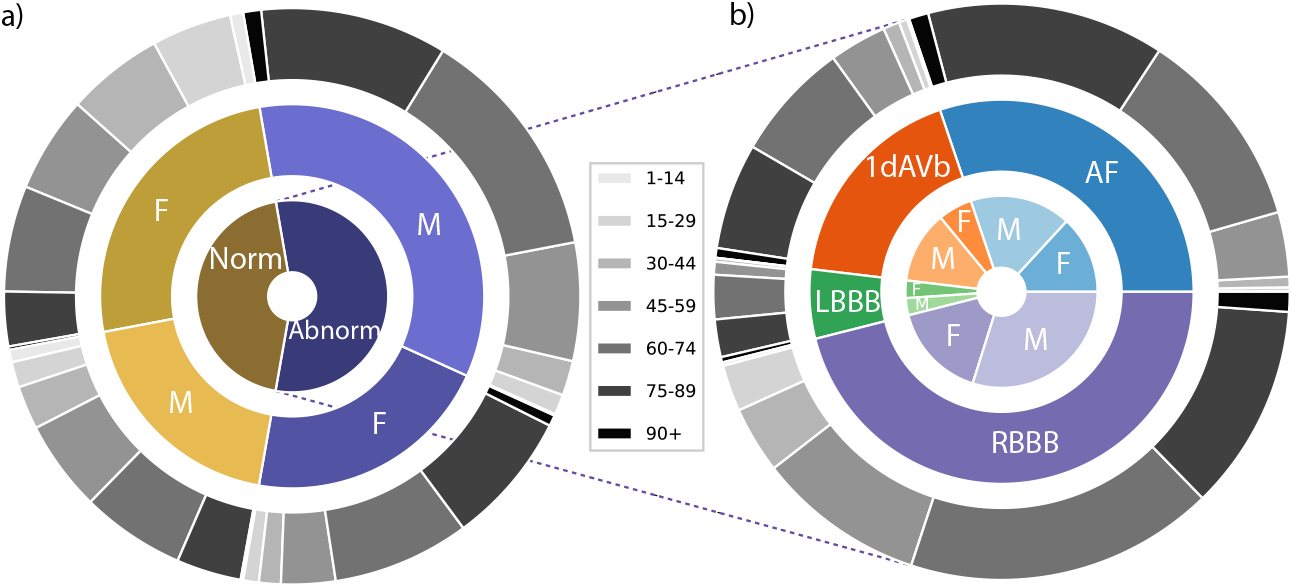
a) Distribution of patients with studied abnormalities versus those with normal ECG readings or other abnormalities in the CPSC dataset. b) Detailed breakdown of specific abnormalities is also shown. Adapted from Huang, Z. et al., J. of Cardiovasc. Trans. Res. (2024) [14]. https://doi.org/10.1007/s12265-024-10504-y. Licensed under CC BY 4.0.

## 5 Experiment

The structure of this section follows the outline presented in the Method. However, since the entire training and *µ*-Training processes are interdependent within the overall framework, we have merged the full training and *µ*-Training into one section. Therefore, we will conduct experiments focusing on the overall training process, while separate experiments will emphasize fine-tuning and its on-device demonstration. Additionally, experiments analyzing the proposed self-distillation will be conducted after those on *µ*-Training and *µ*-Fine-tuning.

### 5.1 Positioning of the *µ*-Training

The training process was configured to run for 300 epochs on the balanced TNMG subset, following the framework outlined in our previous work [8]. However, a key difference in this study is the incorporation of *µ*-Training within the training process.

We conducted a preliminary experiment comparing different positioning strategies to determine the optimal placement of *µ*-Training. The total duration of *µ*-Training was fixed at 100 epochs within the 300-epoch training process. We evaluated three configurations: placing all 100 epochs at the beginning, placing them at the end, and splitting them into two equal 50-epoch segments—one at the beginning and the other at the end. The trained models are evaluated on the TNMG and CPSC datasets. Following the initial training, they undergo *µ*-Fine-Tuning for 50 epochs on CPSC subset before being re-evaluated on both the TNMG and CPSC datasets.

### 5.2 Phase 1 - Training Process (Full Training & *µ*-Training)

The primary objective of this phase is to determine the optimal allocation of epochs between the full-training period and the *µ*-Training period, identifying the most effective breakdown for each stage of the training process. We assume that the full training will be conducted first, followed by *µ*-Training in the experimental setup.

The training process was configured to run for 300 epochs on the balanced TNMG subset, following the framework outlined in our previous work [8]. However, the proposed approach alternates between full training and *µ*-Training phases. Given that the optimal allocation of epochs between these modes is uncertain, we explored various combinations to identify the most effective distribution for maximizing model performance. By training models with different configurations, we systematically compared their performance to determine the best *µ*-Training strategy for achieving optimal results.

During the training phase, the TDSSM operates in two distinct modes: full training and *µ*-Training. Initially, the model is set to full training mode, where all weights are updated using the standard training process. Afterward, the model transitions to *µ*-Training mode, where the encoder and decoder layer weights are frozen, while the middle layer weights remain trainable.

The entire training process spans 300 epochs, consistent with previous model training [8]. To evaluate different configurations, we varied the allocation of epochs between full training and *µ*-Training. Specifically, the full training epochs were set to 0, 50, 100, 150, 200, 250, and 300, with the remaining epochs assigned to *µ*-Training.

### 5.3 Phase 2 - *µ*-Fine-tuning Process

The subsequent fine-tuning experiment was conducted to simulate real-world scenarios where models are adapted to new, personalized data. The CPSC dataset was utilized to evaluate the fine-tuning capabilities of the trained models. In line with previous research indicating that fine-tuning on a balanced dataset yields optimal results [7], a subset of 1,000 samples was extracted from the CPSC dataset. This subset consisted of 200 samples each for the four shared abnormalities between the TNMG and CPSC datasets, along with 200 samples representing healthy data. During this process, the weights in the middle S4D layer are static, while the weights in the encoder and decoder are adjusted to adapt to the new data.

Only the shared abnormalities between the two datasets were considered throughout the fine-tuning process. The differences in abnormalities were carefully managed, with specific measures implemented to ensure that the unused abnormalities in the CPSC dataset did not interfere with the loss calculation. This approach ensured that the finetuning process remained focused and accurate.

### 5.4 Edge Deployment

For additional insight, one model was selected for finetuning directly on the Radxa Zero device, enabling a real-world evaluation of edge device performance compared to Graphics Processing Unit (GPU) based fine-tuning.We conducted experiments using a model trained with a combination of half (150 epochs) full training and half (150 epochs) *µ*-Training. The model was deployed on the proposed Radxa Zero platform for on-device *µ*-Fine-Tuning. The edge fine-tuned model was tested on the test set of TNMG and the entire CPSC dataset. Its performance was compared to a model trained normally for 300 epochs and fine-tuned normally for 50 epochs on the same dataset performed on a GPU.

### 5.6 Model Size Experiments

To evaluate the impact of the proposed method on models of varying sizes, we applied the training process to several variants of the TDSSM model. These variants were generated by adjusting *d*_model_, which controls the model’s width, resulting in models with different sizes and numbers of parameters across layers. The training and fine-tuning processes were repeated for each variant, allowing for a comparative analysis of the results. For this part, we allocated half of the 300 epochs to full training, followed by 150 epochs of *µ*-Training, with *µ*-Fine-tuning set at 50 epochs. The same datasets were used across all trials. We varied *d*_model_ across the values 8, 16, 32, 64, 96, 128, 160, 192, 224, and 256.

### 5.6 Self-Distilled *µ*-Training

The first step we took was to understand the implications of the weighting factor *α*. For our analysis, we have varied the weighting factor *α* of the integrated loss function from 0 to 1 in increments of 0.2 to understand its impact on the overall performance of the model. The parameter *α* controls the proportion of the loss coming from the future-guided distillation loss, with the remaining loss derived from the original loss. The experiment was set up with 150 epochs of full training, followed by 150 epochs of self-distilled *µ*-Training and 50 epochs of *µ*-Fine-tuning.

To demonstrate the effectiveness of our proposed self-guided learning process, we incorporated self-distillation into the *µ*-Training phase while keeping the rest of the process unchanged. Specifically, we used 150 epochs of Full Training, followed by 150 epochs of *µ*-Training and 50 epochs of *µ*-Fine-Tuning. The datasets remained the same across all experiments. The only difference was the inclusion of self-distillation in the *µ*-Training phase. We tested this process on three variants of the model with *d*_model_ values of 96, 128, and 160. The results were then compared with those from the same model variants trained without the self-distillation technique. We will use *α* = 0.8 for the ongoing analysis in this study.

To demonstrate the process of incorporating self-distilled *µ*-Training in the standard training procedure, we implemented a simple training setup where a model with *d*_model_ = 128 undergoes 150 epochs of full training, followed by 75 epochs of self-distilled *µ*-Training on the middle layer, and another 75 epochs of self-distilled *µ*-Training on the encoder and decoder layers. The results were then compared with a standard trained model.

### 5.7 Evaluation Metrics

Evaluation metrics are essential for assessing model performance and ensuring meaningful comparisons. Throughout this work, the detection threshold is set at 0.5. Key metrics include precision, recall, F1-score, and the Area Under the Receiver Operating Characteristic Curve (AUROC), each providing a different perspective on model effectiveness.

Precision (*P*), defined in Eq. 4, measures the proportion of correctly identified positive cases among all predicted positives:

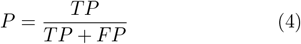

Recall (*R*), given in Eq. 5, evaluates the model’s ability to detect actual positive cases:

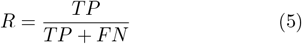

The F1-score (*F*_1_), formulated in Eq. 6, balances precision and recall, offering a single measure of overall performance:

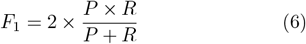

AUROC quantifies the model’s ability to distinguish between positive and negative cases across varying thresholds, making it particularly valuable in binary classification tasks.

These metrics collectively provide a comprehensive evaluation of model performance, particularly in applications like anomaly detection, where distinguishing between classes is critical.

## 6 Results

This section presents the experiments’ results, following the sequence outlined in the previous section.

### 6.1 Positioning of *µ*-Training

We conducted an experiment whose results are summarized in Table 3. The table reports the average AU-ROC—a threshold-independent metric—of the trained and fine-tuned models on both the TNMG and CPSC datasets. In this experiment, the training process comprised a *µ*-Training phase executed over 100 epochs and a full training phase executed over 200 epochs (the order of which may vary), followed by 50 epochs of *µ*-Fine-Tuning.

**Table 3:** Average AUROC for *µ*-Trained and *µ*-Fine-tuned models using various positional strategies—namely, beginning, rear, and sandwich (equally split), evaluated on the TNMG and CPSC datasets.

|  | TNMG | CPSC | CPSC (Fine-tuned) | TNMG (Fine-tuned) |
| --- | --- | --- | --- | --- |
| **Beginning** | 95.41% | 92.07% | 92.37% | 88.81% |
| **Sandwich** | 97.18% | 93.87% | 94.90% | 94.37% |
| **Rear** | 97.51% | 94.30% | 94.51% | 95.47% |

### 6.2 nTraining Process

To evaluate different configurations, we varied the epoch allocation between full training and *µ*-Training, with full training set to 0, 50, 100, 150, 200, 250, and 300 epochs, and the remaining epochs assigned to *µ*-Training. The training process for selected configurations is shown in Fig. 7. The 0 epochs of full training and 300 epochs of *µ*-Training produced unsatisfactory results and is excluded from the chart and analysis.

**Figure 7:**
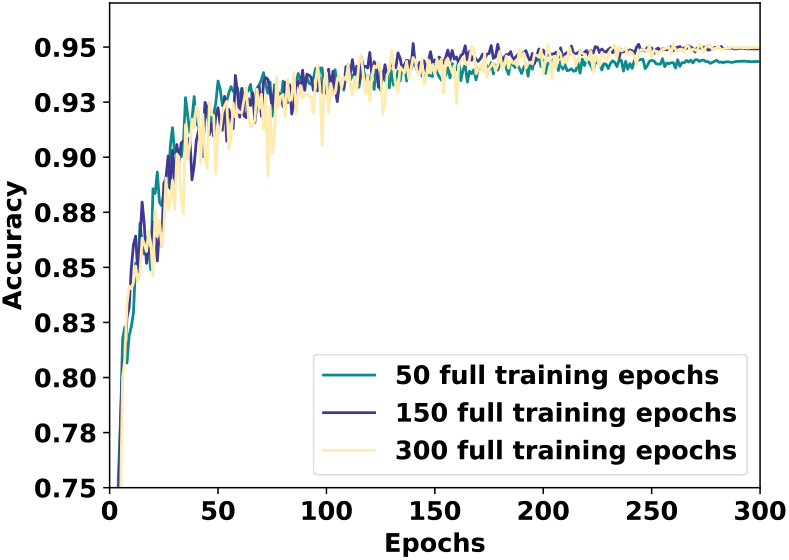
The accuracy evaluation throughout the training process is shown for three selected combinations of full training and *µ*-Training to enhance chart clarity, as including all combinations would make it too cluttered.

After completing the full training and *µ*-Training processes, the model is evaluated using the test dataset from the TNMG subset, which was part of the training process. Additionally, the model’s performance is assessed on the CPSC dataset, serving as unseen data, to evaluate its generalization ability. Fig. 8a) the average F1 score and AUROC are presented, showcasing the model’s performance on the TNMG test set and the CPSC dataset. Table 4 provides a detailed breakdown of the model’s performance on the TNMG test data for each type of abnormality.

**Table 4:**
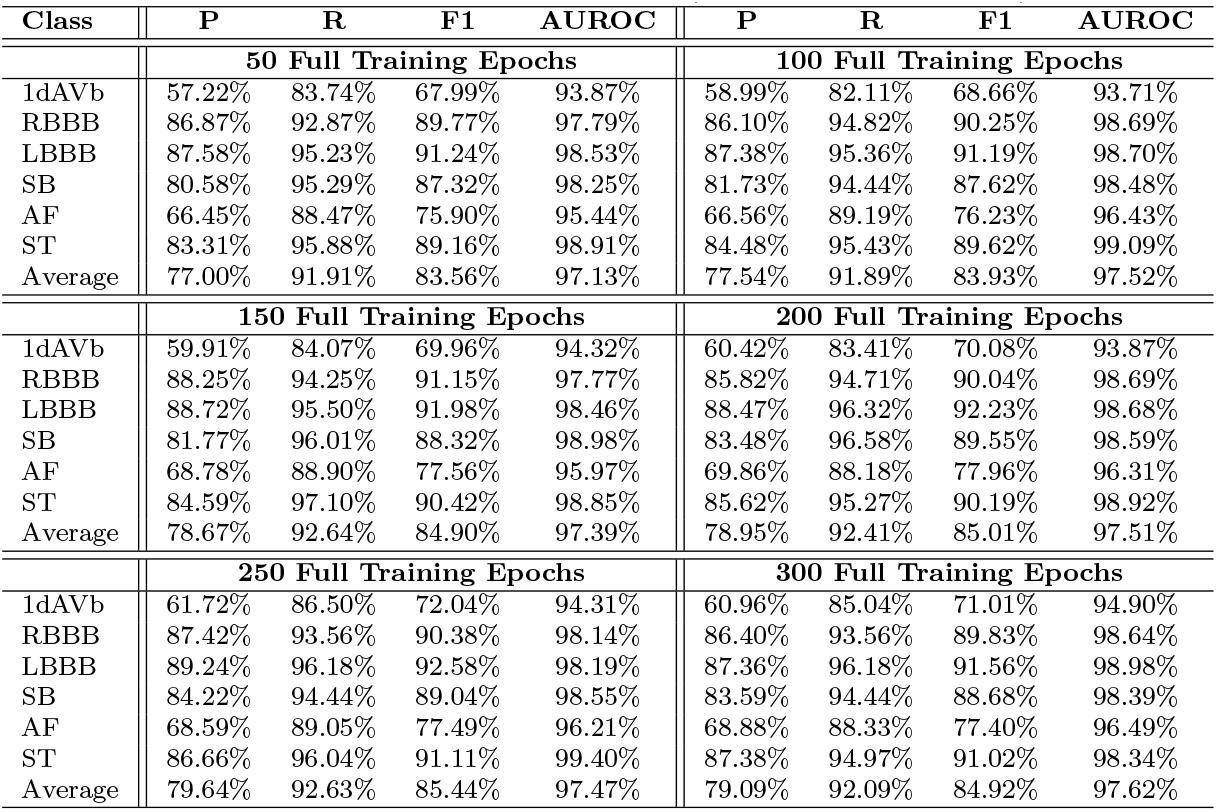
Models Tested on TNMG (P: Precision, R: Recall)

**Figure 8:**
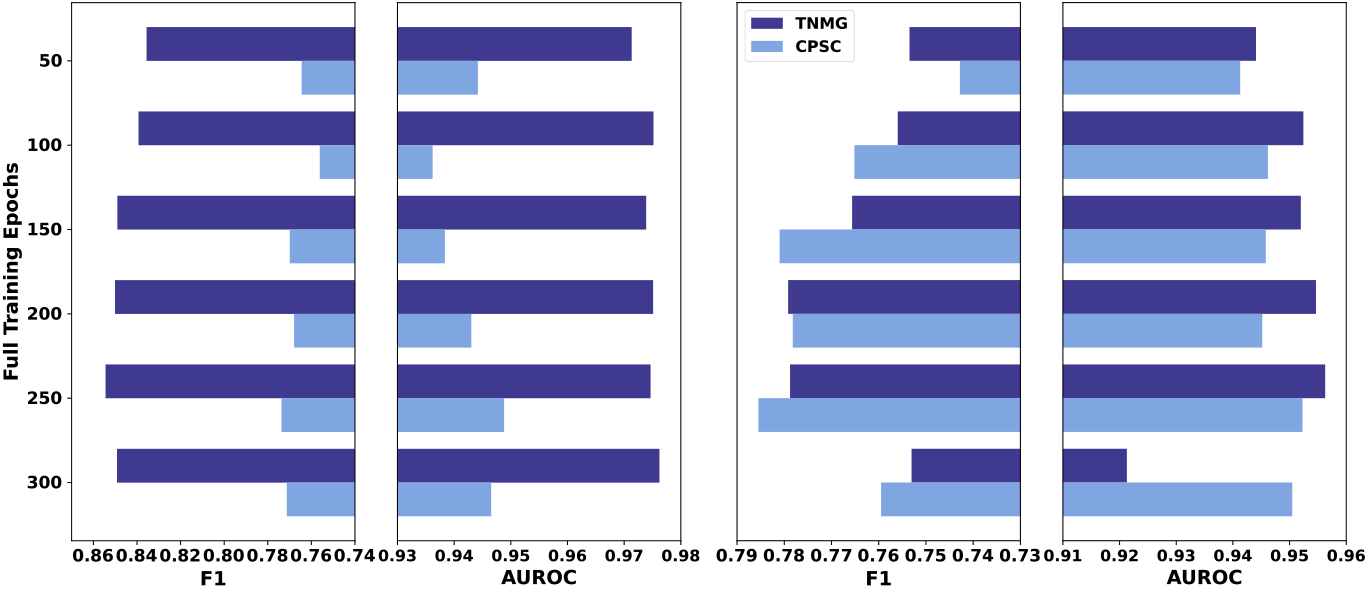
a) Test results of the *µ*-trained models on TNMG and CPSC datasets, with average F1 scores presented on the left and average AUROC scores on the right. The chart’s legend in a) and b) are the same. b) Test results of the *µ*-fine-tuned models on TNMG and CPSC datasets, with average F1 scores on the left and average AUROC scores on the right.

Table 5 presents a detailed analysis of the model’s performance on the CPSC dataset for each type of abnormality. The analysis is limited to the four abnormalities shared between the CPSC and TNMG datasets.

**Table 5:** Models Tested on CPSC (P: Precision, R: Recall)

| Class | P | R | F1 | AUROC | P | R | F1 | AUROC |
| --- | --- | --- | --- | --- | --- | --- | --- | --- |
| 50 Full Training Epochs |  |  |  |  | 100 Full Training Epochs |  |  |  |
| 1dAVb | 62.81% | 86.57% | 72.80% | 94.07% | 61.89% | 84.35% | 71.40% | 93.26% |
| RBBB | 89.48% | 74.69% | 81.42% | 91.08% | 91.44% | 73.67% | 81.60% | 89.76% |
| LBBB | 69.61% | 90.25% | 78.60% | 97.72% | 63.96% | 90.25% | 74.87% | 95.91% |
| AF | 60.47% | 91.97% | 72.97% | 94.80% | 62.14% | 93.28% | 74.59% | 95.55% |
| Average | 70.59% | 85.87% | 76.45% | 94.42% | 69.86% | 85.39% | 75.61% | 93.62% |
| 150 Full Training Epochs |  |  |  |  | 200 Full Training Epochs |  |  |  |
| 1dAVb | 61.14% | 88.92% | 72.46% | 93.64% | 64.95% | 85.73% | 73.91% | 93.18% |
| RBBB | 91.63% | 73.72% | 81.71% | 89.02% | 91.74% | 73.61% | 81.69% | 91.00% |
| LBBB | 66.25% | 88.98% | 75.95% | 96.52% | 65.84% | 89.83% | 75.99% | 97.15% |
| AF | 67.65% | 91.65% | 77.84% | 96.16% | 63.93% | 92.47% | 75.59% | 95.88% |
| Average | 71.67% | 85.82% | 76.99% | 93.84% | 71.62% | 85.41% | 76.79% | 94.30% |
| 250 Full Training Epochs |  |  |  |  | 300 Full Training Epochs |  |  |  |
| 1dAVb | 62.78% | 88.09% | 73.31% | 93.97% | 61.88% | 86.57% | 72.17% | 95.19% |
| RBBB | 92.17% | 74.21% | 82.22% | 92.60% | 91.28% | 75.50% | 82.64% | 90.25% |
| LBBB | 67.08% | 90.68% | 77.12% | 97.18% | 70.06% | 87.29% | 77.74% | 97.21% |
| AF | 65.59% | 92.71% | 76.82% | 95.77% | 64.44% | 92.47% | 75.95% | 95.96% |
| Average | 71.91% | 86.42% | 77.37% | 94.88% | 71.91% | 85.45% | 77.12% | 94.65% |

### 6.3 Fine-tuning Results

Following 50 epochs of fine-tuning, the models are tested again on the CPSC dataset to evaluate changes in performance. The fine-tuning process for various models trained using different combinations of full training and *µ*-Training is illustrated in Fig. 9.

**Figure 9:**
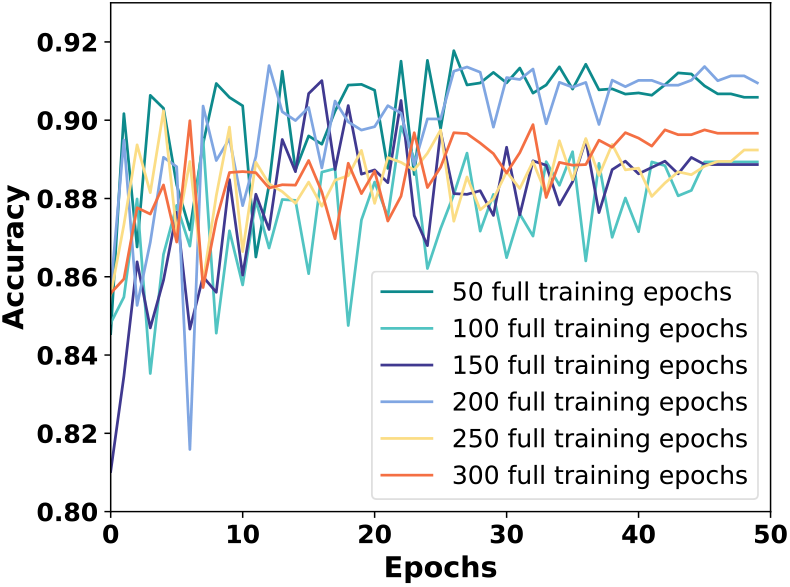
The accuracy evaluation throughout the entire fine-tuning process for various combinations of full training and *µ*-Training.

The fine-tuned models are also tested on the TNMG test set to assess their performance on the original training data. Fig. 8b) summarizes the performance of the finetuned models. Table 6 presents a detailed breakdown of the fine-tuned models’ performance on the TNMG test data, while Table 7 provides a detailed breakdown of their performance on the CPSC dataset.

**Table 6:** *µ*-Fine-tuned Models Tested on TNMG (P: Precision, R: Recall)

| Class | P | R | F1 | AUROC | P | R | F1 | AUROC |
| --- | --- | --- | --- | --- | --- | --- | --- | --- |
| 50 Full Training Epochs |  |  |  |  | 100 Full Training Epochs |  |  |  |
| 1dAVb | 60.57% | 65.20% | 62.80% | 87.84% | 63.43% | 73.33% | 68.02% | 90.62% |
| RBBB | 51.10% | 93.90% | 66.18% | 95.50% | 51.87% | 94.13% | 66.88% | 95.72% |
| LBBB | 90.80% | 90.18% | 90.49% | 97.80% | 91.97% | 92.22% | 92.10% | 97.83% |
| SB | 87.77% | 86.02% | 86.89% | 96.69% | 58.24% | 93.72% | 71.84% | 96.20% |
| AF | 47.22% | 86.89% | 61.19% | 91.40% | 56.16% | 86.74% | 68.18% | 93.62% |
| ST | 79.05% | 90.85% | 84.54% | 97.21% | 83.15% | 90.24% | 86.55% | 97.46% |
| Average | 69.42% | 85.51% | 75.35% | 94.41% | 67.47% | 88.40% | 75.60% | 95.24% |
| 150 Full Training Epochs |  |  |  |  | 200 Full Training Epochs |  |  |  |
| 1dAVb | 69.13% | 65.53% | 67.28% | 90.55% | 64.89% | 69.43% | 67.09% | 90.62% |
| RBBB | 49.15% | 93.67% | 64.48% | 95.13% | 52.61% | 91.60% | 66.83% | 94.35% |
| LBBB | 92.39% | 91.13% | 91.76% | 97.53% | 90.22% | 90.59% | 90.40% | 97.08% |
| SB | 74.66% | 93.72% | 83.11% | 98.38% | 73.73% | 94.86% | 82.97% | 97.27% |
| AF | 60.74% | 84.73% | 70.76% | 94.03% | 59.59% | 88.62% | 71.26% | 95.27% |
| ST | 74.20% | 91.62% | 81.99% | 95.56% | 85.00% | 93.29% | 88.95% | 98.21% |
| Average | 70.05% | 86.73% | 76.56% | 95.20% | 71.01% | 88.07% | 77.92% | 95.46% |
| 250 Full Training Epochs |  |  |  |  | 300 Full Training Epochs |  |  |  |
| 1dAVb | 78.21% | 65.37% | 71.21% | 90.85% | 65.98% | 67.48% | 66.72% | 91.31% |
| RBBB | 55.98% | 91.60% | 69.49% | 94.68% | 53.72% | 92.98% | 68.10% | 94.33% |
| LBBB | 90.82% | 91.81% | 91.32% | 97.61% | 85.63% | 94.27% | 89.74% | 98.58% |
| SB | 72.65% | 93.58% | 81.80% | 98.28% | 94.32% | 63.91% | 76.19% | 81.32% |
| AF | 56.73% | 88.62% | 69.18% | 94.11% | 52.63% | 88.04% | 65.88% | 92.94% |
| ST | 78.45% | 91.01% | 84.26% | 98.23% | 88.63% | 82.01% | 85.19% | 94.30% |
| Average | 72.14% | 87.00% | 77.88% | 95.63% | 73.48% | 81.45% | 75.30% | 92.13% |

**Table 7:**
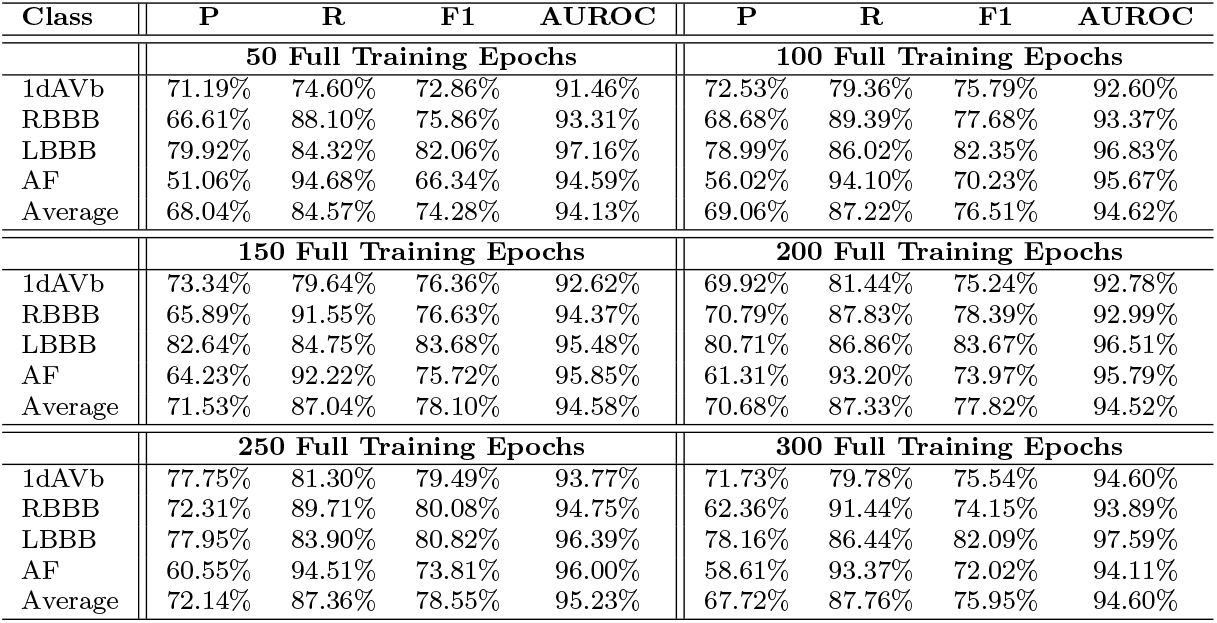
*µ*-Fine-tuned Models Tested on CPSC (P: Precision, R: Recall)

| Class | P | R | F1 | AUROC | P | R | F1 | AUROC |
| --- | --- | --- | --- | --- | --- | --- | --- | --- |
| 50 Full Training Epochs |  |  |  |  | 100 Full Training Epochs |  |  |  |
| 1dAVb | 71.19% | 74.60% | 72.86% | 91.46% | 72.53% | 79.36% | 75.79% | 92.60% |
| RBBB | 66.61% | 88.10% | 75.86% | 93.31% | 68.68% | 89.39% | 77.68% | 93.37% |
| LBBB | 79.92% | 84.32% | 82.06% | 97.16% | 78.99% | 86.02% | 82.35% | 96.83% |
| AF | 51.06% | 94.68% | 66.34% | 94.59% | 56.02% | 94.10% | 70.23% | 95.67% |
| Average | 68.04% | 84.57% | 74.28% | 94.13% | 69.06% | 87.22% | 76.51% | 94.62% |
| 150 Full Training Epochs |  |  |  |  | 200 Full Training Epochs |  |  |  |
| 1dAVb | 73.34% | 79.64% | 76.36% | 92.62% | 69.92% | 81.44% | 75.24% | 92.78% |
| RBBB | 65.89% | 91.55% | 76.63% | 94.37% | 70.79% | 87.83% | 78.39% | 92.99% |
| LBBB | 82.64% | 84.75% | 83.68% | 95.48% | 80.71% | 86.86% | 83.67% | 96.51% |
| AF | 64.23% | 92.22% | 75.72% | 95.85% | 61.31% | 93.20% | 73.97% | 95.79% |
| Average | 71.53% | 87.04% | 78.10% | 94.58% | 70.68% | 87.33% | 77.82% | 94.52% |
| 250 Full Training Epochs |  |  |  |  | 300 Full Training Epochs |  |  |  |
| 1dAVb | 77.75% | 81.30% | 79.49% | 93.77% | 71.73% | 79.78% | 75.54% | 94.60% |
| RBBB | 72.31% | 89.71% | 80.08% | 94.75% | 62.36% | 91.44% | 74.15% | 93.89% |
| LBBB | 77.95% | 83.90% | 80.82% | 96.39% | 78.16% | 86.44% | 82.09% | 97.59% |
| AF | 60.55% | 94.51% | 73.81% | 96.00% | 58.61% | 93.37% | 72.02% | 94.11% |
| Average | 72.14% | 87.36% | 78.55% | 95.23% | 67.72% | 87.76% | 75.95% | 94.60% |

### 6.4 Edge Deployment

During the edge *µ*-Fine-tuning process, the system utilized a total of 3 GB of RAM, including 1.2 GB when idle, ensuring that the resources required for fine-tuning remained below 2 GB. The edge fine-tuned model’s performance on the TNMG and CPSC datasets is compared to a GPU-trained model in Table 8.

**Table 8:** Comparison of *µ*-Training and on-device *µ*-Fine-Tuning vs. Normal Training and Fine-Tuning Process (P: Precision, R: Recall)

| Class | P | R | F1 | AUROC | P | R | F1 | AUROC |
| --- | --- | --- | --- | --- | --- | --- | --- | --- |
| Tested on TNMG Dataset |  |  |  |  |  |  |  |  |
| μ-Trained and μ-Fine-tuned | | | | | Standard Trained and Fine-tuned | | | |
| 1dAVb | 68.93% | 67.80% | 68.36% | 91.14% | 75.95% | 48.78% | 59.41% | 86.35% |
| RBBB | 47.97% | 92.41% | 63.15% | 93.66% | 53.71% | 93.33% | 68.18% | 94.40% |
| LBBB | 91.83% | 91.95% | 91.88% | 97.99% | 87.78% | 92.09% | 89.88% | 98.10% |
| SB | 71.41% | 97.29% | 82.37% | 98.50% | 94.23% | 74.61% | 83.28% | 90.64% |
| AF | 60.52% | 86.60% | 71.25% | 94.02% | 60.31% | 84.29% | 70.31% | 93.12% |
| ST | 91.16% | 84.91% | 87.92% | 93.93% | 54.62% | 98.17% | 70.19% | 98.36% |
| Average | 71.97% | 86.83% | 77.49% | 94.87% | 71.10% | 81.88% | 73.54% | 93.50% |
| Tested on CPSC Dataset |  |  |  |  |  |  |  |  |
| μ-Trained and μ-Fine-tuned | | | | | Standard Trained and Fine-tuned | | | |
| 1dAVb | 72.14% | 82.13% | 76.81% | 93.18% | 80.45% | 64.96% | 71.88% | 90.90% |
| RBBB | 64.95% | 91.71% | 76.04% | 94.37% | 70.04% | 90.90% | 79.12% | 94.35% |
| LBBB | 80.71% | 86.86% | 83.67% | 95.70% | 79.28% | 84.32% | 81.72% | 96.24% |
| AF | 59.62% | 92.87% | 72.62% | 95.48% | 58.57% | 91.48% | 71.42% | 94.67% |
| Average | 69.36% | 88.39% | 77.29% | 94.68% | 72.09% | 82.92% | 76.04% | 94.04% |

### 6.5 Size Matters

We created a series of models by adjusting *d*_model_, resulting in models of different sizes. When *µ*-Training is applied, the ratio of trainable parameters to the total number of parameters varies with the model size. Each model undergoes 150 epochs of full training, followed by 150 epochs of *µ*-Training. The models are then evaluated on the TNMG dataset and tested on the CPSC dataset to assess generalization. After completing the training process, they are further *µ*-Fine-Tuned for 50 epochs and re-evaluated on TNMG and CPSC. The results are summarized in Fig. 10.

**Figure 10:**
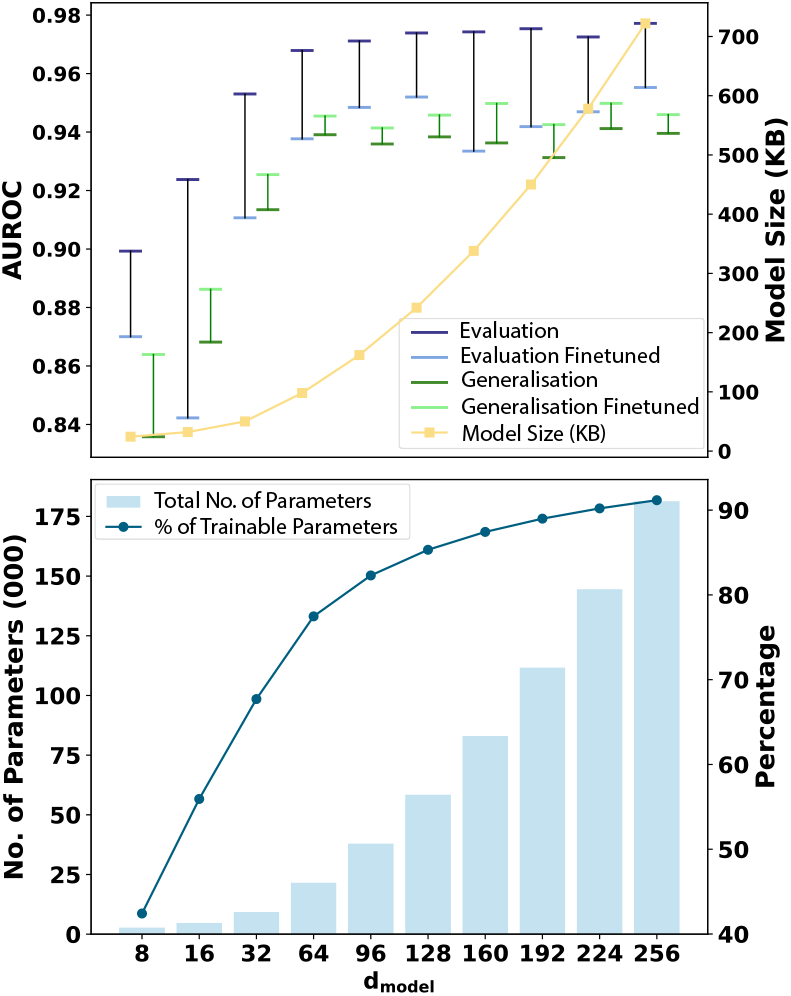
a) illustrates the change in average AUROC for models with varying *d*_model_ before and after *µ*-Fine-Tuning, evaluated on TNMG for evaluation and CPSC for generalization. The primary y-axis represents AU-ROC, while the secondary y-axis indicates the model sizes in KB. b) The primary y-axis displays the total number of parameters for models with different *d*_model_ values, while the secondary y-axis represents the percentage of trainable parameters during the *µ*-Training stage.

### 6.6 Self-distilled *µ*-Training

While we have described the weighting factor *α* for the proposed loss function in the self-distilled *µ*-Training process, we further investigated its impact on performance by varying *α*. Fig. 11 illustrates the effect of different *α* values on the model’s performance.

**Figure 11:**
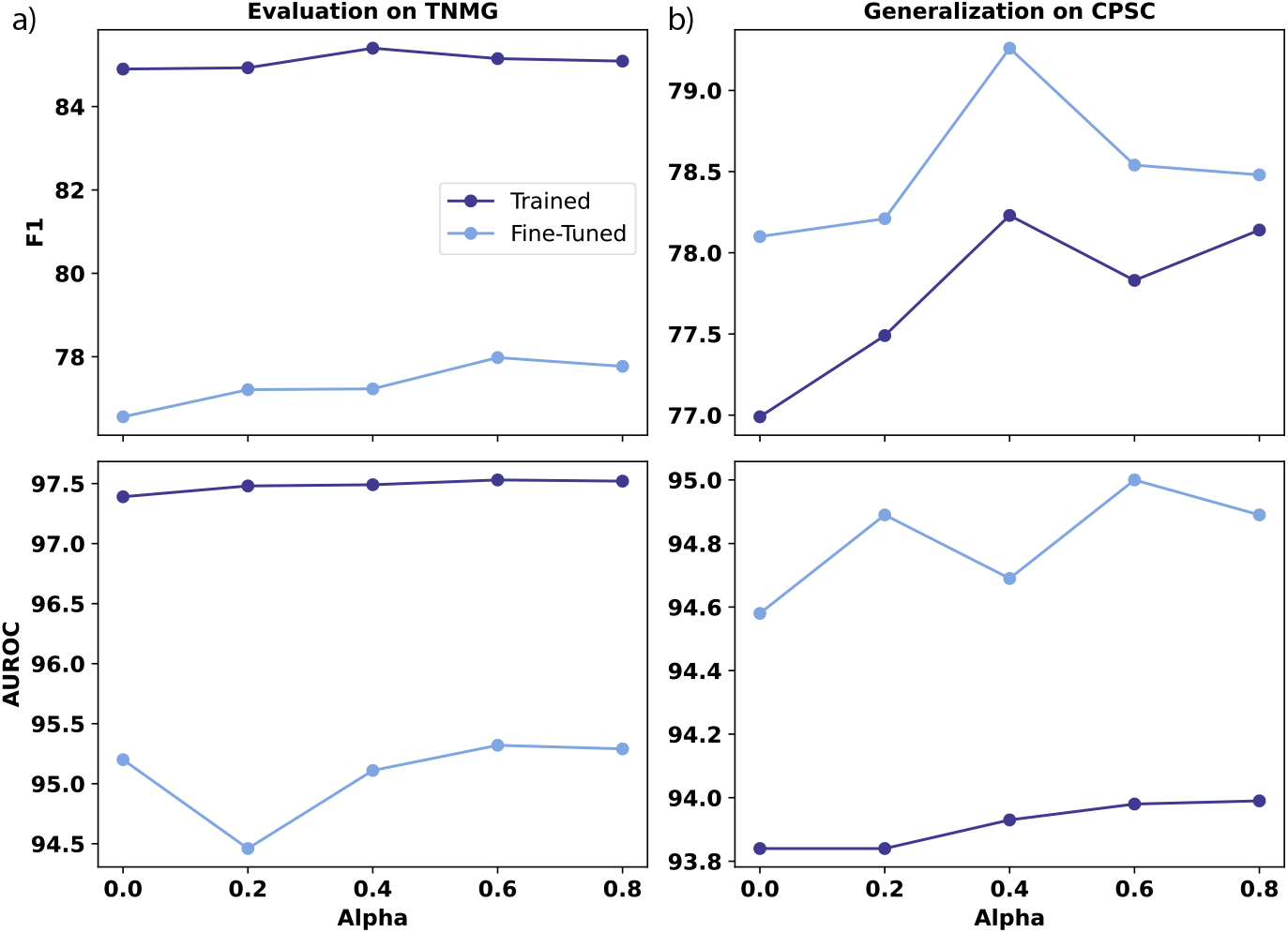
The figure illustrates: a) the average F1 and AUROC of the model trained with 150 epochs of full training followed by 150 epochs of future-guided self-distilled *µ*-training, using varied weighting factors (*α*) in the training data, TNMG, along with the corresponding results after 50 epochs of *µ*-fine-tuning; and b) the model’s performance on the generalization dataset, CPSC, and after fine-tuning with CPSC to evaluate its performance with different values of *α*.

To evaluate our proposed self-distillation incorporated into *µ*-Training, we tested it on three models with *d*_model_ values of 96, 128, and 160 separately. These models previously demonstrated stable performance in earlier results. The experiment followed the same procedure as before, with the only difference being self-distillation integration during the *µ*-Training stage. The results, before and after *µ*-Fine-Tuning, on TNMG for evaluation and CPSC for generalization, are summarized in Fig. 12. The model with *d*_model_ = 128 underwent a complete self-distilled *µ*-Training process, beginning with 150 epochs of full training, followed by 75 epochs of self-distilled *µ*-Training on the middle layer, and another 75 epochs of self-distilled *µ*-Training on the encoder and decoder layers. We also applied *µ*-Fine-tuning. The average F1 score and AUROC are compared to the same model, which underwent standard training and fine-tuning process as illustrated in Fig. 13.

**Figure 12:**
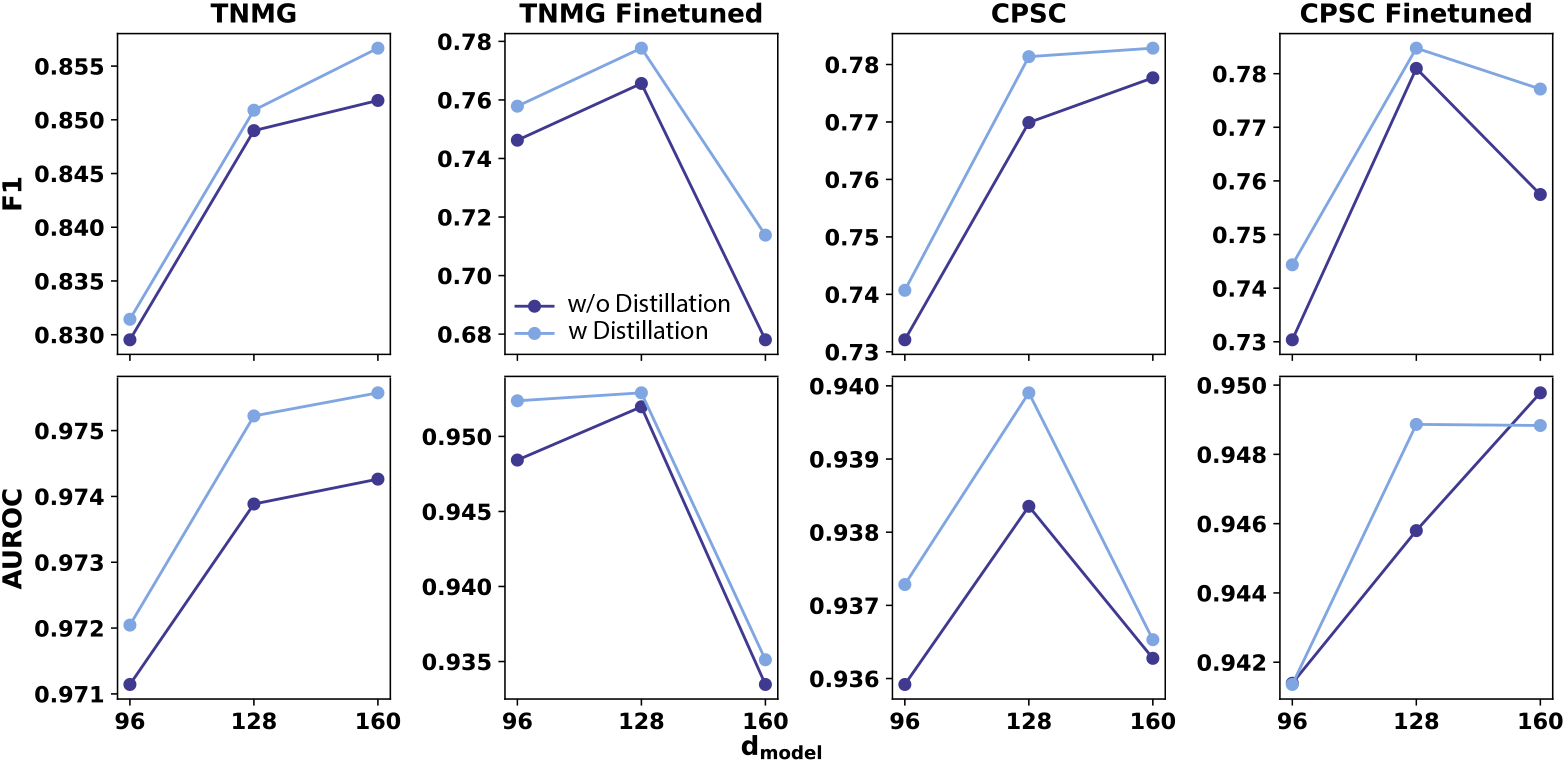
Average F1 score (Top) and AUROC (Bottom) of models with *d*_model_ values of 96, 128, and 160, comparing the proposed self-distilled *µ*-Training to models trained without self-distillation. Performance is evaluated separately on TNMG for assessment and CPSC for generalization, followed by re-evaluation on TNMG and CPSC after *µ*-Fine-Tuning.

**Figure 13:**
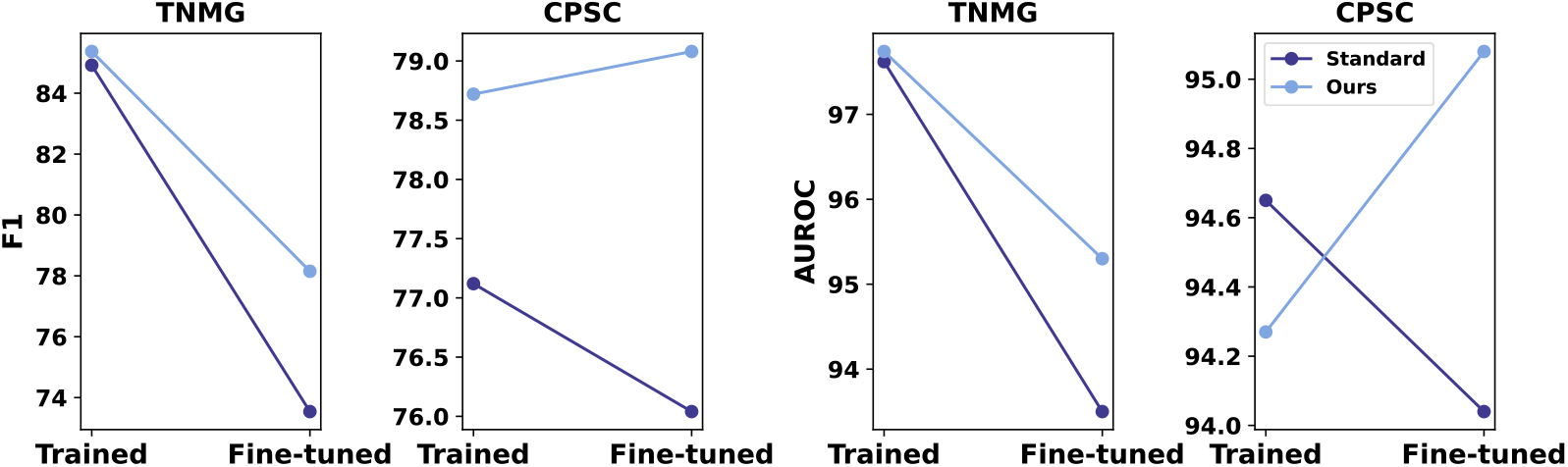
Average F1 score (left) and AUROC (right) for a model with *d*_model_ = 128 on TNMG and CPSC datasets, completely trained using our proposed self-distilled *µ*-Training and *µ*-Fine-tuning methods, compared to a model trained and fine-tuned via standard techniques.

## 7 Discussion

The discussion will be structured to follow the order in which the results are presented in Section 6.

### 7.1 Impact of Positioning

The placement of *µ*-Training within the overall training process affects model performance. Our results indicate that placing *µ*-Training at the end yields the best performance on both the training dataset (TNMG) and the generalization dataset (CPSC) when the model is completely trained. Although a sandwich configuration—allocating half of *µ*-Training at the beginning and half at the end—slightly outperforms the end-only strategy on CPSC after the *µ*-Fine-tuning process, the end-only positioning offers more stable results with minimal performance drop on TNMG after fine-tuning. While further investigation into the sandwich approach could provide additional insights and potentially enhance the method, for this study, we have chosen to position *µ*-Training at the end.

### 7.2 Impact on Training

We experimented with various combinations of full training and *µ*-Training. The key observation is that the combined approach performs nearly as well as standard full training, where the model completes 300 epochs of full training. Fig. 7, 8a), and Table 4 show that incorporating *µ*-Training alongside full training does not compromise the model’s performance on the test data from the training dataset. This is particularly evident in metrics such as the average F1 score and AUROC, which reflect the balanced performance of the models. Notably, the optimal performance is achieved when the model undergoes more full training than *µ*-Training.

When the trained model was tested on the CPSC dataset, which serves as unseen data, its behavior closely mirrored the observations made during testing on the training dataset’s test set. The models’ performance did not significantly decline compared to those trained with the standard 300 epochs. As shown in Fig. 8a) and Table 5, the best performance was observed when the model underwent more full training than *µ*-Training.

Overall, the key observation is that models trained using a combination of full training and *µ*-Training maintain comparable performance to those trained using the standard process, in both in-sample and out-of-sample tests.

### 7.3 Impact on *µ*-Fine-Tuning

The impact of *µ*-Fine-tuning yields intriguing results. After fine-tuning, the model was tested again on both the test set of the trained TNMG dataset and the CPSC dataset. As shown in Fig. 9 and Fig. 8b), models trained through the standard training process alone did not perform as optimally as those that underwent a combination of full training and *µ*-Training. Models trained with both methods demonstrated better performance on the CPSC and the TNMG dataset’s test set, indicating that they retain knowledge of the original data.

A closer examination of Table 6 and Table 7 provides additional support for the earlier inference. The standard trained model does not perform as optimally as the models trained using the combined approach, particularly in terms of F1 and AUROC, which assess the balanced performance of the model. These findings confirm that combining full training with *µ*-Training outperforms the standard training process when the models undergo *µ*-Fine-tuning with limited resource requirements on local edge devices.

Furthermore, the proposed method helps mitigate forgetting more effectively than models that do not utilize it. This demonstrates its strong performance in preventing catastrophic forgetting, as models trained with the proposed method retain more of the original data they were initially trained on after fine-tuning on new data.

### 7.4 Edge Deployment

To validate our proposed method, we deployed a model that is partially full trained and partially *µ*-Trained onto the Radxa Zero platform to test the effectiveness of *µ*-Fine-tuning. The experimental results are summarized in Table 8. Also included in the table are the performance metrics of the model that underwent traditional training and fine-tuning on a GPU. The results are noteworthy, as the models that underwent full training, *µ*-Training, and *µ*-Fine-tuning at the edge outperform the standard trained and fine-tuned model on the CPSC dataset. Interestingly, these models also perform better when tested on the training TNMG dataset. This demonstrates that our proposed method not only achieves superior results when computational power and memory are constrained through *µ*-training and fine-tuning, but also outperforms traditional training and fine-tuning processes.

### 7.5 Impact by Size

As the model size increases, its performance improves and stabilizes around 128. Additionally, larger models exhibit better generation performance than smaller ones. Among all models, the one with *d*_*model*_ = 160 demon-strates the best generalization improvement, which suggests that it achieves an optimal balance between model complexity and generalization ability.

Additionally, as model size increases, the trainable parameters in the *µ*-Training process grows, leading to a higher trainable percentage. However, this comes at the cost of increased computational and memory requirements, which may not always be justified by marginal improvements in performance. The results highlight the importance of selecting an optimal model size to balance efficiency and effectiveness, with *d*_*model*_ = 128 or 160 emerging as a strong candidate for maximizing generalization while maintaining manageable resource demands. This also provides valuable insights into model design, helping ensure that *µ*-Training and *µ*-Fine-Tuning achieve optimal performance. The ideal ratio of trainable parameters to total parameters appears to fall between 15% and 20%, offering a good balance between performance improvement and computational efficiency. These findings are particularly useful for the design of *µ*-Trainer and, ultimately, the foundational model [7].

### 7.6 Impact of Self-Distillation

In terms of *α*, the model trained with *α* = 0.8 demonstrates stable performance. However, after *µ*-fine-tuning, some other values of *α* slightly outperform it in certain metrics. Notably, performance declined when testing *α* = 1, where the loss function relies entirely on future-guided self-distillation. This suggests that with *α* = 1, training is driven only by matching the gradients from the top and bottom layers, which account for a small portion of the model’s overall weights. As a result, this guidance alone is insufficient for effective learning, highlighting the need for some dependence on ground truth labels, especially in this three-layer setting where the middle layer holds the majority of the model’s weights.

Across all tested models, the model incorporating self-distillation consistently demonstrates improvements in average F1 score and AUROC, except one outlier: the average AUROC of the model with *d*_model_ of 160 in the post-fine-tuned generalization.

For *d*_*model*_ = 96, 128, and 160, self-distillation increases average F1 scores and AUROC in both evaluation and fine-tuned settings. Notably, the model with *d*_*model*_ = 128 exhibits the best improvement in generalization performance with self-distillation, suggesting its effectiveness in enhancing its ability to maintain high performance across different datasets.

Integrating the novel self-distillation technique in *µ*-Training holds significant potential for broader applications beyond medical and TinyML fields. This new approach can be adapted to existing traditional training processes, enabling the calibration of models for enhanced performance and improved generalization.

We conducted a simple experiment to demonstrate its effectiveness, as outlined in earlier sections. Our experiment shows that our model outperformed the standard-trained version on TNMG (higher average F1 and AU-ROC) and achieved higher average F1 on CPSC despite slightly lower average AUROC. After *µ*-Fine-tuning, it exceeded the standard fine-tuned model on both datasets across both metrics.

### 7.7 Comparison with Leading Methods

Table 9 presents a qualitative comparison of lightweight and personalized edge learning methods. It highlights the key differences in approach, personalization, training efficiency, security & privacy, accuracy trade-offs, and datasets used. Notably, our method, which employs *µ*-Training with self-distillation and *µ*-Fine-Tuning, achieves high training efficiency and privacy protection while maintaining minimal accuracy loss. Unlike other frameworks that rely on FL or knowledge caching, our approach processes data locally, reducing security risks. The evaluation spans diverse datasets, including CPSC and TNMG.

**Table 9:** Comparison of Lightweight and Personalized Edge Learning Methods.

| Ref | Approach | Personalization | Training Efficiency | Security & Privacy | Accuracy Trade-Off | Dataset Used |
| --- | --- | --- | --- | --- | --- | --- |
| [35] | Adaptive pruning with personalized FL | Yes | Medium | Medium | Moderate accuracy loss | FEMNIST |
| [36] | Sparse training with task adaptation (TinyTrain) | Yes | Medium | Low | Moderate accuracy loss | CIFAR-10, Tiny ImageNet |
| [37] | Decentralized FL framework (EdgeFL) | Yes | Medium | Medium | Minimal accuracy loss | MNIST, CIFAR-10 |
| [38] | Lightweight FL framework (FLight) | No | Medium | Medium | Minimal accuracy loss | MINST, SIFAR |
| [39] | Federated learning with knowledge caching (FedCache 2.0) | No | Medium | Medium | Minimal accuracy loss | CIFAR-10, CINIC-10, CIFAR-100 |
| [40] | Semi-supervised personalized FL (SemiPFL) | Yes | Medium | Medium | Minimal accuracy loss | MOBIACT, WISDM, HAR-UCI, HHAR, PAMAP2 |
| **Our work** | μ-Training and μ-Fine-Tuning with self-distillation | Yes | High | High | Minimal accuracy loss | CPSC, TNMG |

FL has been widely regarded as a privacy-preserving method, as it updates model gradients without transmitting users’ raw data to a central server. However, recent research has demonstrated that original data can be reconstructed from these gradient updates, revealing potential privacy risks [4, 5]. As a result, the security of FL is not as robust as previously assumed. FL may require higher computational requirements due to its reliance on decentralized data processing and the need for frequent full model updates [41]. This approach also leads to higher communication overhead, as each participant must transmit large amounts of model updates to a central server, which may result in communication bottlenecks. Additionally, the accumulation of updates from numerous devices can cause an overload, further reducing the efficiency of the learning process. This combination of server dependencies, communication overload, and the complexity of full model updates makes FL more challenging in real-time decision-making and efficient use of resources [42].

### 7.8 Privacy & Security

While on-device training significantly enhances privacy by eliminating the need to transmit sensitive data, it also demands careful consideration of potential vulnerabilities. For example, resource-constrained edge devices may be particularly susceptible to adversarial attacks that exploit their limited local data diversity, leading to model misbehavior [43]. Moreover, isolating training data can constrain the model’s exposure to diverse scenarios, potentially reducing its generalizability and robustness against sophisticated attacks.

By integrating Future-Guided Self-Distilled *µ*-Training with *µ*-Fine-Tuning, our approach effectively balances computational efficiency, performance, and privacy. Processing personal data entirely on edge devices ensures that sensitive information is never transmitted or exposed to external systems, thereby fundamentally reducing the attack surface compared to traditional FL frameworks [4]. Additionally, Future-Guided Self-Distilled *µ*-Training significantly improves the generalizability of the pre-trained model while mitigating catastrophic forgetting through the subsequent application of *µ*-Fine-Tuning. This process enhances the model’s resilience to sophisticated adversarial attacks by ensuring that learning remains robust over time. Although this decentralized strategy may sacrifice some benefits derived from aggregated learning, such as exposure to a wider variety of data, it ultimately provides superior privacy and personalization compared to conventional FL methods.

## 8 Limitations and Future Works

Despite the promising results of the proposed methodology, several limitations warrant discussion. First, while the TDSSM model demonstrates lightweight characteristics and high efficiency, its applicability may be constrained to specific types of bio-signal data, such as ECG signals. The generalizability of the proposed technique to other bio-signal modalities or medical applications remains uncertain. Future work could extend the experimental framework to include additional datasets, such as those for electroencephalogram (EEG) or photoplethysmography (PPG) signals, to validate the versatility of the approach.

Another limitation lies in the computational resources and hardware employed for on-device fine-tuning. While the Radxa Zero was chosen for its resource-efficient design, the experimental results may not fully capture the constraints imposed by other edge devices with differing architectures or lower processing capabilities. Testing the methodology across a broader range of hardware platforms could offer deeper insights into its practicality and scalability for diverse deployment environments.

The *µ*-Training and *µ*-Fine-Tuning processes, although effective in improving the model’s adaptability and reducing computational overhead, rely heavily on optimal hyperparameter configurations, such as the allocation of epochs between phases. The lack of a standardized approach for determining these configurations introduces a degree of subjectivity that may impact reproducibility. Future research could explore automated hyperparameter optimization techniques.

The design of the TDSSM model was adopted from prior work [8], and it was not explicitly designed to accommodate the proposed methodology. This inherent limitation suggests that the efficiency of the proposed *µ*-Training and *µ*-Fine-Tuning techniques may not be optimal, as the design of the encoder and decoder layers, along with the distribution of weights across layers, could significantly influence the effectiveness of the strategy. Future work could involve redesigning the model architecture to optimize the number of weights and layer structure to better align with the proposed methodology. Such refinements could enhance both the computational efficiency and overall performance of the technique, providing a more robust framework for bio-signal data analysis.

The proposed method may also apply to other tinyML models beyond the TDSSM tested in this study. This technique can be extended to other application domains, particularly in conjunction with the growing adoption of IoT. Such expansion could enable broader real-world applications, subject to further testing and experimentation.

While the proposed self-distillation has shown great promise, further testing is still needed, particularly regarding the design of the loss function, including the weighing ratio and specific distillation loss for multi-binary classification models. That being said, the self-distilled *µ*-Training approach shows potential for broader applications beyond TinyML. A more calibrated training strategy, focusing on specific layers, could further improve model performance.

The proposed method has also shown promise in combating catastrophic forgetting by focusing learning on specific parts of the model, allowing other parts to adapt to new information while preserving previously learned knowledge. While effective, the method can be further refined to better address scenarios where mitigating catastrophic forgetting is crucial.

## 9 Conclusion

In conclusion, this study introduces a novel approach to training and fine-tuning deep learning models for bio-signal analysis, using the TDSSM model as a demonstrative example. By combining full training with *µ*-Training, we demonstrate that the model can achieve comparable performance to standard training while significantly reducing computational overhead. Including *µ*-Fine-Tuning further enhances the model’s ability to adapt to new data with minimal resource consumption, making it well-suited for edge deployment in resource-constrained environments. The results from both the TNMG and CPSC datasets support the effectiveness of the combined approach, highlighting its potential for generalization and performance improvement. Our experiments also show that deploying the model on the Radxa Zero platform for edge *µ*-Fine-Tuning yields superior results compared to traditional training and fine-tuning processes, offering a compelling solution for bio-signal processing. We have also integrated a novel self-distillation technique into the *µ*-Training process, further enhancing the performance of the proposed method. The self-distilled *µ*-Training shows promising potential for applications beyond medical fields and TinyML. The proposed method also significantly reduces catastrophic forgetting after being *µ*-fine-tuned on new data.

However, future work should investigate the applicability of this method to other bio-signal types, explore automated hyperparameter optimization techniques, and test the approach on a broader range of hardware platforms to further assess its scalability and robustness. With the potential integration of tinyML and IoT, the proposed method could also benefit other models with broader applications outside the medical domain. A carefully calibrated training method based on the self-distilled *µ*-Training concept could also benefit the broader deep learning field.

## Data Availability

This research paper uses the public CPSC dataset but acknowledges that the TNMG dataset is private and requires permission from its owner for access.

## 10 Acknowledgment

Zhaojing Huang gratefully acknowledges the support the Australian Government’s Research Training Program (RTP) provides.

## 11 Availability of Code

The code is available at the following link: https://github.com/NeuroSyd/uTraining. Please note that specific terms, conditions, or usage restrictions may apply.

## 12 Availability of Data

This research paper uses the publicly available CPSC dataset. The TNMG dataset is accessible through a pathway for interested readers to obtain the data through its custodians via an agreement, which requires permission for access.

## 14 Ethics Statement

The authors confirm that all data used in this paper (see Section 12) are publicly accessible, either directly or through a request to data custodians. They comply with their relevant laws and institutional guidelines and have been approved by the appropriate institutional committees. Our use of the datasets mentioned in this paper fully complies with the terms and conditions of their use.

## References

[1] Bing Shi and Yiming Wu. Task offloading and resource allocation strategies among multiple edge servers. IEEE Internet of Things Journal, 11(8):14647–14656, 2023.

[2] Nduma N Basil, Solomon Ambe, Chukwuyem Ekhator, and Ekokobe Fonkem. Health records database and inherent security concerns: A review of the literature. Cureus, 14(10), 2022.

[3] Mansoor Ali, Faisal Naeem, Muhammad Tariq, and Georges Kaddoum. Federated learning for privacy preservation in smart healthcare systems: A comprehensive survey. IEEE Journal of Biomedical and Health Informatics, 27(2):778–789, 2022.

[4] Ligeng Zhu, Zhijian Liu, and Song Han. Deep leakage from gradients. Advances in Neural Information Processing Systems, 32, 2019.

[5] Franziska Boenisch, Adam Dziedzic, Roei Schuster, Ali Shahin Shamsabadi, Ilia Shumailov, and Nicolas Papernot. Reconstructing individual data points in federated learning hardened with differential privacy and secure aggregation. In IEEE 8th European Symposium on Security and Privacy (EuroS&P), pages 241–257, 2023.

[6] An Wang, Jiong Yu, Danqi Liu, and Yong Cui. A measurement study for transmission energy on mobile device. In Proceedings of 2012 2nd International Conference on Computer Science and Network Technology, pages 708–714. IEEE, 2012.

[7] Zhaojing Huang, Leping Yu, Luis Fernando Herbozo Contreras, Kamran Eshraghian, Nhan Duy Truong, Armin Nikpour, and Omid Kavehei. Advancing privacy-aware machine learning on sensitive data via edge-based continual µ-training for personalized large models. Machine Learning: Science and Technology, 6(1):015025, 2025.

[8] Zhaojing Huang, Wing Hang Leung, Jiashuo Cui, Leping Yu, Luis Fernando Herbozo Contreras, Nhan Duy Truong, Armin Nikpour, and Omid Kavehei. Cardiac abnormality detection with a tiny diagonal state space model based on sequential liquid neural processing unit. APL Machine Learning, 2(2), 2024.

[9] Polychronis E Dilaveris and Harold L Kennedy. Silent atrial fibrillation: epidemiology, diagnosis, and clinical impact. Clinical Cardiology, 40(6):413– 418, 2017.

[10] Ayodeji Oseni, Nour Moustafa, Helge Janicke, Peng Liu, Zahir Tari, and Athanasios Vasilakos. Security and privacy for artificial intelligence: Opportunities and challenges. arXiv preprint 2102.04661, 2021.

[11] Lachit Dutta and Swapna Bharali. TinyML meets IoT: A comprehensive survey. Internet of Things, 16:100461, 2021.

[12] Zhaojing Huang, Luis Fernando Herbozo Contreras, Leping Yu, Nhan Duy Truong, Armin Nikpour, and Omid Kavehei. S4D-ECG: A shallow state-of-the-art model for cardiac abnormality classification. Cardiovascular Engineering and Technology, pages 1–12, 2024.

[13] Albert Gu, Karan Goel, Ankit Gupta, and Christopher Ré. On the parameterization and initialization of diagonal state space models. Advances in Neural Information Processing Systems, 35:35971– 35983, 2022.

[14] Zhaojing Huang, Luis Fernando Herbozo Contreras, Wing Hang Leung, Leping Yu, Nhan Duy Truong, Armin Nikpour, and Omid Kavehei. Efficient edge-AI models for robust ECG abnormality detection on resource-constrained hardware. Journal of Cardiovascular Translational Research, pages 1–14, 2024.

[15] Mathias Lechner, Ramin Hasani, Alexander Amini, Thomas A Henzinger, Daniela Rus, and Radu Grosu. Neural circuit policies enabling auditable autonomy. Nature Machine Intelligence, 2(10):642– 652, 2020.

[16] Zhaojing Huang, Wing Hang Leung, Leping Yu, Luis Fernando Herbozo Contreras, Ziyao Zhang, Nhan Duy Truong, Armin Nikpour, and Omid Kavehei. On-device edge-learning for cardiac abnormality detection using a bio-inspired and spiking shallow network. APL Machine Learning, 2(2), 2024.

[17] Ziming Liu, Yixuan Wang, Sachin Vaidya, Fabian Ruehle, James Halverson, Marin Soljačić, Thomas Y. Hou, and Max Tegmark. KAN: Kolmogorov-arnold networks, 2024.

[18] Zhaojing Huang, Jiashuo Cui, Leping Yu, Luis Fernando Herbozo Contreras, and Omid Kavehei. Abnormality detection in time-series bio-signals using kolmogorov-arnold networks for resource-constrained devices. medRxiv, pages 2024–06, 2024.

[19] Li Li, Yuxi Fan, Mike Tse, and Kuo-Yi Lin. A review of applications in federated learning. Computers & Industrial Engineering, 149:106854, 2020.

[20] Massimo Merenda, Carlo Porcaro, and Demetrio Iero. Edge machine learning for AI-enabled IoT devices: A review. Sensors, 20(9):2533, 2020.

[21] Tommaso Furlanello, Zachary Lipton, Michael Tschannen, Laurent Itti, and Anima Anandkumar. Born again neural networks. In International conference on machine learning, pages 1607–1616. PMLR, 2018.

[22] Zhilu Zhang and Mert Sabuncu. Self-distillation as instance-specific label smoothing. Advances in Neural Information Processing Systems, 33:2184–2195, 2020.

[23] Geoffrey Hinton, Oriol Vinyals, and Jeff Dean. Distilling the knowledge in a neural network. arXiv preprint 1503.02531, 2015.

[24] Adriana Romero, Nicolas Ballas, Samira Ebrahimi Kahou, Antoine Chassang, Carlo Gatta, and Yoshua Bengio. Fitnets: Hints for thin deep nets. arXiv preprint 1412.6550, 2014.

[25] Linfeng Zhang, Jiebo Song, Anni Gao, Jingwei Chen, Chenglong Bao, and Kaisheng Ma. Be your own teacher: Improve the performance of convolutional neural networks via self distillation. In Proceedings of the IEEE/CVF international conference on computer vision, pages 3713–3722, 2019.

[26] Antônio H Ribeiro, Manoel Horta Ribeiro, Gabriela MM Paixão Derick M Oliveira, Paulo R Gomes, Jéssica A Canazart, Milton PS Ferreira, Carl R Andersson, Peter W Macfarlane, Wagner Meira Jr, et al. Automatic diagnosis of the 12lead ECG using a deep neural network. Nature Communications, 11(1):1760, 2020.

[27] Feifei Liu, Chengyu Liu, Lina Zhao, Xiangyu Zhang, Xiaoling Wu, Xiaoyan Xu, Yulin Liu, Caiyun Ma, Shoushui Wei, Zhiqiang He, et al. An open access database for evaluating the algorithms of electrocardiogram rhythm and morphology abnormality detection. Journal of Medical Imaging and Health Informatics, 8(7):1368–1373, 2018.

[28] Euan A Ashley and Josef Niebauer. Arrhythmia. In Cardiology explained. Remedica, 2004.

[29] Kenneth A Mayuga, Artur Fedorowski, Fabrizio Ricci, Rakesh Gopinathannair, Jonathan Walter Dukes, Christopher Gibbons, Peter Hanna, Dan Sorajja, Mina Chung, David Benditt, et al. Sinus tachycardia: a multidisciplinary expert focused review. Circulation: Arrhythmia and Electrophysiology, 15(9):e007960, 2022.

[30] Sunjeet Sidhu and Joseph E Marine. Evaluating and managing bradycardia. Trends in cardiovascular medicine, 30(5):265–272, 2020.

[31] Nicholas Y Tan, Chance M Witt, Jae K Oh, and Yong-Mei Cha. Left bundle branch block: current and future perspectives. Circulation: Arrhythmia and Electrophysiology, 13(4):e008239, 2020.

[32] AK Agarwal and P Venugopalan. Right bundle branch block: varying electrocardiographic patterns.: Aetiological correlation, mechanisms and electrophysiology. International journal of cardiology, 71(1):33–39, 1999.

[33] Yoshihiro Tanaka, Ravi B Patel, Hayato Tada, Kenshi Hayashi, Masayuki Takamura, Masa-aki Kawashiri, and Philip Greenland. First-degree atrioventricular block is associated with incident atrial fibrillation in the japanese general population aged ≥ 40 years. Circulation, 142(Suppl 3):A13015– A13015, 2020.

[34] Zhaojing Huang, Sarisha MacLachlan, Leping Yu, Luis Fernando Herbozo Contreras, Nhan Duy Truong, Antonio Horta Ribeiro, and Omid Kavehei. Generalization challenges in electrocardiogram deep learning: insights from dataset characteristics and attention mechanism. Future Cardiology, pages 1–12, 2024.

[35] Yueying Zhou, Gaoxiang Duan, Tianchen Qiu, Lin Zhang, Li Tian, Xiaoying Zheng, and Yongxin Zhu. Personalized federated learning incorporating adaptive model pruning at the edge. Electronics, 13(9):1738, 2024.

[36] Young D Kwon, Rui Li, Stylianos I Venieris, Jagmohan Chauhan, Nicholas D Lane, and Cecilia Mascolo. Tinytrain: resource-aware task-adaptive sparse training of dnns at the data-scarce edge. arXiv preprint 2307.09988, 2023.

[37] Hongyi Zhang, Jan Bosch, and Helena Holmström Olsson. Edgefl: A lightweight decentralized federated learning framework. In 2024 IEEE 48th Annual Computers, Software, and Applications Conference (COMPSAC), pages 556–561. IEEE, 2024.

[38] Wuji Zhu, Mohammad Goudarzi, and Rajkumar Buyya. Flight: A lightweight federated learning framework in edge and fog computing. Software: Practice and Experience, 54(5):813–841, 2024.

[39] Quyang Pan, Sheng Sun, Zhiyuan Wu, Yuwei Wang, Min Liu, Bo Gao, and Jingyuan Wang. Fed-cache 2.0: Federated edge learning with knowledge caching and dataset distillation. arXiv preprint 2405.13378, 2024.

[40] Arvin Tashakori, Wenwen Zhang, Z Jane Wang, and Peyman Servati. Semipfl: Personalized semisupervised federated learning framework for edge intelligence. IEEE Internet of Things Journal, 10(10):9161–9176, 2023.

[41] Laizhong Cui, Xiaoxin Su, Yipeng Zhou, and Yi Pan. Slashing communication traffic in federated learning by transmitting clustered model updates. IEEE Journal on Selected Areas in Communications, 39(8):2572–2589, 2021.

[42] Goran Saman Nariman and Hozan Khalid Hamarashid. Communication overhead reduction in federated learning: a review. International Journal of Data Science and Analytics, pages 1–32, 2024.

[43] Raissa Souza, Pauline Mouches, Matthias Wilms, Anup Tuladhar, Sönke Langner, and Nils D Forkert. An analysis of the effects of limited training data in distributed learning scenarios for brain age prediction. Journal of the American Medical Informatics Association, 30(1):112–119, 2023.

